# PICASA: Decomposing patient heterogeneity of single-cell cancer data by cross-attention neural networks

**DOI:** 10.1101/2025.06.04.25328900

**Authors:** Sishir Subedi, Yongjin P Park

## Abstract

**Motivation:** Gene expression variation in cancer cells is attributed to many inherited and environmental factors, including genetic variants and cellular landscapes. Decomposing different sources of information is intractable with single-cell RNA-seq alone. However, we show that our new approach, PICASA, can split them with the help of multiple patients, assuming that cell types are widely shared and genetic effects are specifically present in a particular patient. Our approach, based on a cross-attention neural network, was applied to diverse cancer types to identify cell types and patient-specific genetic effects in transcriptomic data. The method highlights residual expressions, excluding cell types, which can implicate patient-specific disease mechanisms.

**Results:** PICASA effectively decomposes cell type commonality and the residual patient-specific signature in three different complex cancer datasets, including breast, ovarian, and lung cancers. Unlike many existing distance-based batch adjustment methods (unable to recover cell-type-specific generative models), PICASA learns transferrable gene/feature embedding coordinates and cell-type-specific gene-gene interaction patterns as an attention layer. We also demonstrate that many cancer patient-specific signatures captured by PICASA are deemed somatic and genetic, such as copy number variants (CNV).

**Availability and implementation:** PICASA source code is available at https://github.com/causalpathlab/picasa. Additionally, to ensure reproducibility, data analysis and figure generation code is available at https://github.com/causalpathlab/picasa/tree/main/picasa_reproducibility.

## Introduction

Single-cell genomic techniques have emerged as powerful tools for unbiased profiling of cancer cells. These advancements provide a deeper understanding of cancer heterogeneity and the complex regulatory networks [1]. Traditional single-cell analysis methods often assume that the data generation process in cancer is largely similar to single-cell data obtained from healthy or reference tissues. However, single-cell transcriptomic data derived from multiple cancer patients show unique characteristics [2] from the corresponding reference tissues and cell types of non-cancer samples. Here, we recognize that extracting cellular commonality shared across many patient-derived samples, in contrast to patient-specific (potentially genetic) patterns [3], is a key question to be addressed in high-dimensional single-cell cancer data analysis.

### Related work

Large-scale single-cell datasets provide unbiased resources to study intrinsic biological mechanisms and patient-specific environmental/genetic factors. However, integrating datasets over tens of patients poses significant computational challenges. The traditional approach focuses on reconciling low-dimensional latent representations (cellular embedding) across different batches and data modalities [4] in order to result in unbiased results in downstream clustering and cell-type-specific differential gene expression (DGE) analysis. Several notable methods build on graph-based data integration framework, where cell-cell interaction graphs come from k-nearest neighbour search, including the first version of a mutual nearest neighbour method [5], Scanorama [6], BBKNN (batch-balancing k-nearest neighbour) [7], Harmony [8], and the weighted nearest neighbour method tailored for multimodal integration [9]. The performance of these distance graph-based methods arguably relies more on quality latent representation learning than on the algorithmic matching and adjustment procedures.

An atlas-scale benchmark study focusing on data integration emphasizes that model-based approaches, such as a variational autoencoder model [10], yield generally richer and robust latent feature space [11]. The notable examples include SCVI [12] and scANVI [13], where we can directly incorporate batch information and covariates in the generative model to remove potential biases present in single-cell data. However, as the research community has gradually become cautious about the risks of directly manipulating latent space to produce visually appealing two-dimensional scatter plots [14], demands for objective and systematic evaluation criteria have grown. A recent study warns that potentially harmful artifacts could be seen among several model-based methods [15]. Model parameters can be easily unidentifiable if a deep learning algorithm has to rely on noisy observations, while not encouraging the models to pick up invariant features.

More recently, several approaches have been proposed for variance decomposition in multi-donor single-cell data integration problems. CellANOVA [16] employs a linear mixed effect modelling approach to separating out condition-specific and/or batch effects from common (fixed) effects estimated by existing batch adjustment methods, such as Harmony [8] trained on SVD features. Factorization methods, GEDI [17] and Lemur [18], seek to isolate donor-specific or condition-specific signals, which are often marginalized in traditional data integration methods. The idea is to include two *a priori* independent components in the matrix factorization models: gene-to-latent dictionary parameters and latent-to-cell latent variables. A deep “fair” autodencoder model [19], such as ScANVI [13] and Biolord [20], also modifies latent-to-cell variables to disentangle multiple sources of variation in latent space. We find a similarity with our previous work, `deltaTopic` [21], modelling distribution shifts. However, environmental or conditional effects considered in the previous work are intrinsically different from patient-specific effects that we normally observe in cancer data. ConstrastiveVI [22] is designed to address similar questions, hoping that shared and condition-specific latent dimensions are statistically independent with the knowledge of control/background cells, similar to RUV (removing unwanted variation) methods based on control genes and samples [23]. In practice, we normally have little knowledge of control cells in heterogeneous cancer data; hence, the method is not always applicable.

### Our contributions

We propose a deep learning approach, PICASA, short for Partitioning Inter-patient Cellular Attributes by Shared Attention networks, to systematically dissect multiple sources of variation embedded in cancer transcriptomic data. Our novelty lies in a tailored neural network architecture and training algorithm that can effectively tackle practical challenges practitioners frequently encounter in multi-donor cancer single-cell data analysis. We hypothesize that intrinsic cellular effects are invariantly manifested in the majority of patients/donors/samples. Built on the success of foundation models in single-cell data [24–27], we show that ubiquitous cell type signatures can be robustly redeemed by training cell pairs as if clinicians understand tumour heterogeneity in light of matched normal samples. Having the common cellular contexts established, we show that patient or donor-specific factors can be better understood. For each of the three different cancer examples, including breast [28], ovarian [29], and lung cancer [30], we show that a cell type commonality and the residual patient-specific signature models are effectively decomposed by PICASA while being markedly independent of each other.

## Methods

### Datasets, study design and preprocessing

We obtained two groups of publicly available datasets-one group contains simulated and real data for integration benchmark experiments, and the other group contains cancer patient single-cell data from different cancer types (Supplementary S1). We conducted a series of standard single-cell data preprocessing and quality-control steps. First, we filtered out mitochondrial and spike genes along with genes detected in less than three cells. Then, we normalized cells by a fixed sequencing depth, i.e., 10^4^, followed by log normalization and high variable gene selection. Throughout all the experiments, we used 2,000 highly variable genes for the initial unsupervised learning for the commonality model and brought all the features back for the subsequent analysis with quality control procedures recommended in `Scanpy` library [31].

#### Lung cancer

The lung cancer dataset [30] was retrieved from Gene Expression Omnibus (GEO) with the accession number GSE148071. The dataset consists of 42 tissue biopsy samples, ranging from stage III to IV non-small cell lung cancer (NSCLC). We selected 23 patient samples that contain at least 1000 cells and constructed a dataset with 72,514 total cells, including 26,077 genes.

#### Ovarian cancer

The high-grade serous ovarian cancer (HGSOC) dataset, collected from eleven patients before and after chemotherapy [29] was retrieved from Gene Expression Omnibus GSE165897. All the patients were homogeneously treated with post-neoadjuvant chemotherapy (NACT) as recommended for inoperable patients with poor prognosis.

#### Breast cancer

The breast cancer single-cell dataset of multiple subtypes [28], including *ER*+, *HER2*+, and Triple Negative Breast Cancer (TNBC), is publicly available at GEO with the accession number GSE176078. Of the 26 patients, we retained cells collected from 21 patients, of which we found at least 1,000 cells.

#### Normal pancreas single-cell data

The total dataset consists of 14,890 cells, each of which contains 2,000 genes, across eight different sequencing batches. We downloaded them from Seuret data integration tutorial, https://satijalab.org/seurat/archive/v3.2/integration.html, but each of them is also publicly available from NCBI GEO GSE81076, GSE85241, GSE86469, and GSE84133, and EBI Array Express: E-MTAB-5061.

### Overview of PICASA

We design PICASA in order to decompose high-dimensional single-cell gene expression vectors. PICASA first seeks to identify inherent cell type patterns robustly shared across multiple patients and then characterizes patient-specific gene expression patterns by contrasting them with the common signatures. Our premise is that gene expression patterns derived from cell type signatures are widely present across multiple cancer patients and matched normal samples, as they reflect inherent tissue-of-origin characteristics. However, we argue that somatic mutations and environmental factors are manifested in a patient-specific manner.

PICASA consists of two types of neural network models (Supplementary S5) with the following purposes:

1. **A commonality network shared across patients**: Briefly, we designed a specialized neural network architecture that recognizes the commonality across nearest neighbour pairs of cells between matched patients and trained it with a contrastive learning method [32,33] (see below for details). The model learns patient-invariant latent representations (Fig. 1) and gene-gene interaction networks of each cell type or cell state (e.g., Fig. 2B).
2. **A patient-specific network**: Provided that this recognition network can faithfully encode latent states of cell states robustly shared across patients, we capture the remaining patient-specific (unique) gene expression signals with the second neural network built on a transitional variational autoencoder model [34]. Here, we used negative binomial autoencoder model (`scvi`) [12] while conditioning on a part of the latent representation learned from the commonality network.

**Fig. 1.**
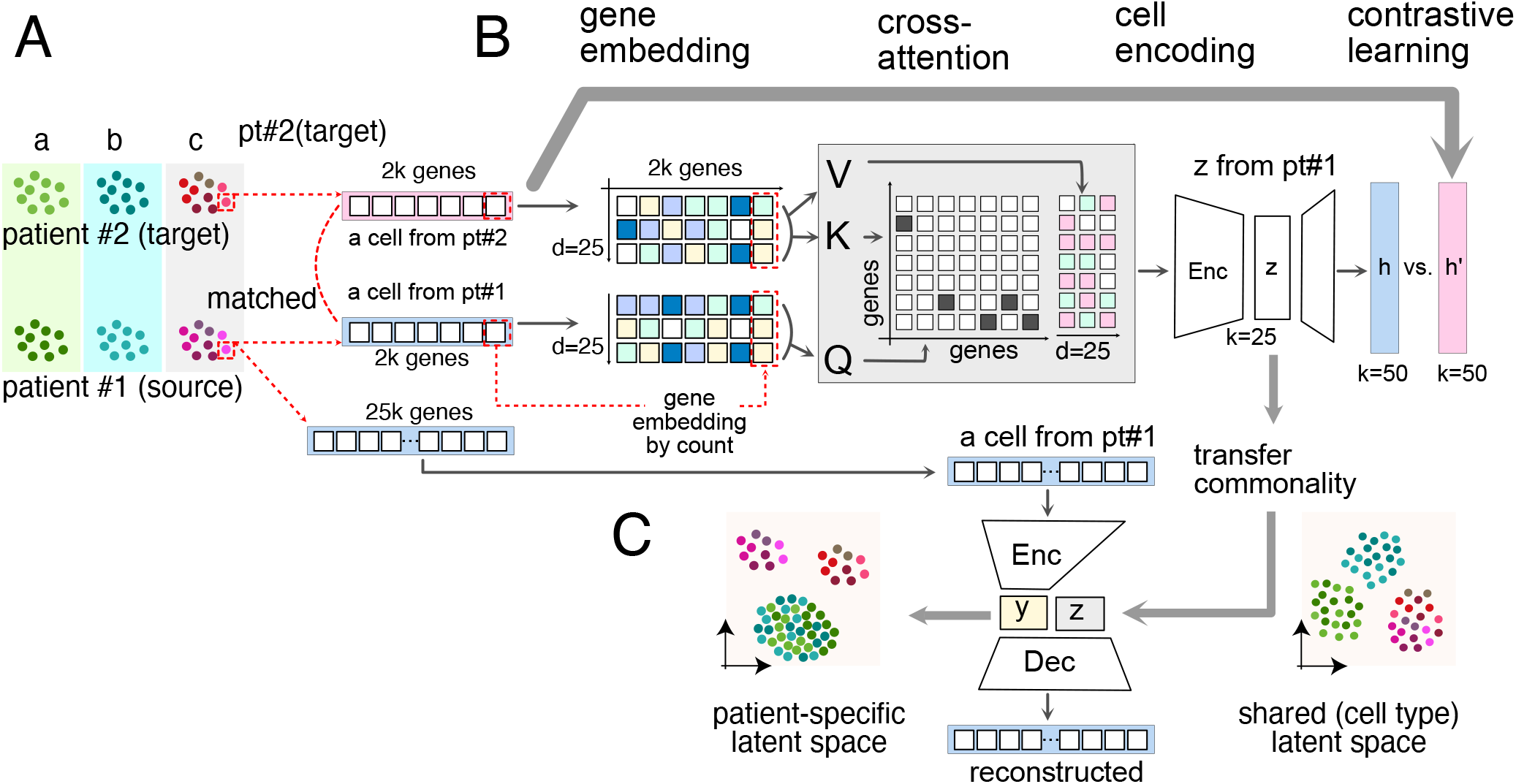
Overview of PICASA. PICASA model consists of two networks: A commonality network for identifying shared biological cell state representation across patients and a patient-specific network for capturing patient-specific effects. **(A)** The approximate neighbor search method estimates pairwise single-cell data from two patients. **(B)** The cell-pair data is embedded in high-dimension at the gene level to share gene interactions and projected onto common latent space, capturing shared biological cell states using contrastive learning. **(C)** A normalized gene expression matrix and pre-trained common latent space by the commonality network are used in the patient-specific network consisting of encoder-decoder architecture. The model learns patient-specific representation from the original expression data not present in the shared common representation.

**Fig. 2.**
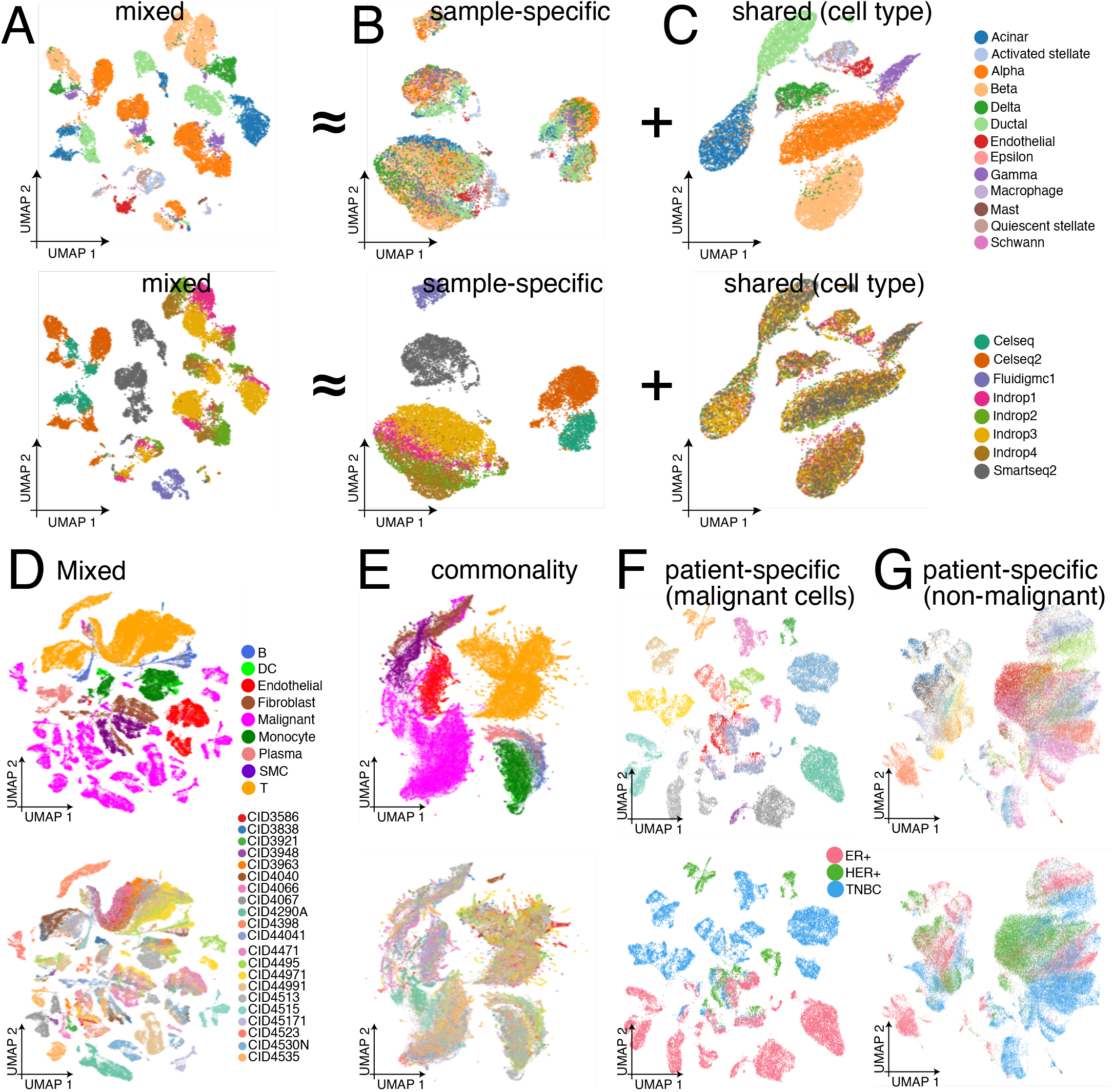
PICASA disentangle sample-specific and sample-invariant effects in pancreas and breast cancer data. PICASA captures shared cell type and sequencing platform-specific batch effects in normal pancreas data. UMAP of **(A)** mixed and decomposed **(B)** common and **(C)** unique representations (upper panel is cell type label and lower panel is sample label). PICASA captures shared cell type and hormone receptor-driven patient-specific effects in breast cancer. UMAP of **(D)** mixed and decomposed **(E)** common representations. UMAP of **(F)** cancer and **(G)** non-cancer cells from unique representation space learned by the patient-specific network (upper panel is cell type label and lower panel is sample label).

#### Single-cell data

The model takes a gene expression count matrix, *X*, with *n* rows (cells) and *m* columns (genes); each element takes non-negative value, namely *X*_*ig*_ ∈ ℝ_≥0_, for a cell *i* and a gene *g*. Some studies share a count matrix of normalized values, so the values can be a rational number. Since such a count matrix was collected on each patient *p*, we specifically mark each count matrix with its patient of origin, namely, *X*^(*p*)^. Likewise, we denote a row vector of a cell *i* by **x**_*i*_ with the patient of origin *p* and use 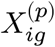 for each element of a gene *g* in a cell *i* observed in a patient *p*.

#### Learning commonality with cross-attention mechanisms between patients

The commonality network architectures consist of three components (Fig. 1B): (1) gene embedding, (2) cross-attention between a pair of cells derived from different patients, and (3) cell encoding. All the parameters of these components are estimated by a contrastive learning algorithm, which puts generative training to a classification problem [32,33].

##### (1) Gene embedding space

The goal of gene embedding is to locate a gene expression value in semantically identifiable locations. Roughly speaking, a traditional gene expression mapping locates values in the same real line, but our embedding scheme systematically allocates each gene and its expression values into fine-grained positions. We may consider gene embedding as an automatic quantization step. For each cell, we first take log-transformation, namely 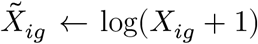, followed by the depth normalization to 10,000, so that 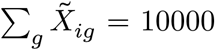. Without loss of too much generality, we partition the transformed and normalized expression values into 3,000 bins, meaning that our expression vocabulary size is kept at that level. An embedding layer allocates a cell vector 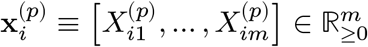 to enlarged feature space ℝ^*m*×*d*^, where *d* is 25 in all the models. We used `torch.nn.Embedding` implemented in `pyTorch` library.

We may introduce an additional embedding scheme based on gene symbols, as `scGPT` [25] does, if we want our embedding layer to be universally applicable across different data sets and studies. Since we assume commonalities across cancer cells are captured by the top 2,000 highly expressed genes, and the downstream attention layers always see the same order of genes, such a gene symbol-based embedding does not play a significant role. Each gene’s identity is implicitly identifiable by its position.

##### (2) Cross-attention layers to recognize the commonality

*How can we represent common cellular mechanisms?* Gene regulatory mechanisms are arguably best represented by multiple snapshots of gene-gene interaction networks. In a large language model (LLM), attention mechanisms have been shown to be powerful enough to generate realistic and meaningful sentences by simply tracking which pairs of words (tokens) are associated within a sentence and a paragraph [35], and so can be the meaning of a cell (gene expression tokens). In order to capture invariant gene interaction networks, we train a pair of gene token vectors (cells) sampled across different patients.

For instance, our cross-attention layer seeks to associate expression tokens of a cell *i* derived from a patient *p* with another, yet similar, cell *j* of a different patient *q*, and vice versa (Supplementary S3). More precisely, we have the embedded genes/tokens, *E*^(*i*)^ ∈ ℝ^*m*×*d*^, for the cell *i*, where *m* = 2000, the number of genes (fixed order), and *d*=25, the embedding dimension, and *E*^(*j*)^ for the counterpart cell *j* (Fig. 1A-B). In order to feed the embedding vector for each gene 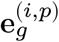 into the scaled-dot product attention layer, we construct query, key, and value matrices with the dense model parameter matrix *W*:

- Query: 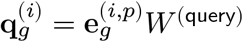,
- Key: 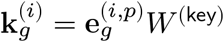,
- Value: 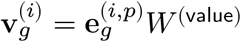,

Having two sets of query, key, and value tensors on both sides of cells, *i* and *j*, we can synthesize a cross-attention vector for a gene *g* from the cell *i* to *j* as if we are mapping a source cell *i* to a target cell *j*’s context (Fig. 1B):

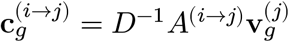

where an unscaled cross-attention matrix *A* takes each element,

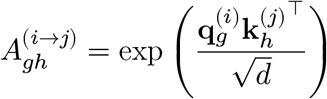

with the normalizer matrix,

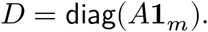

These cross-attention intermediate representations, *C*^(*i*→*j*)^ ∈ ℝ^*m*×*d*^, are then pooled across the embedding dimension, and constitute a cell-level score vector ∈ ℝ^*m*^:

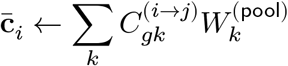

Using two fully-connected (FC) layers (the first with 100 and the second with 25 units), we can then map the pooled gene vector into a low-dimensional latent space:

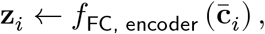

just as in a typical single-cell variational autoencoder model, such as `scvi` [12].

For donors *p* and *q* matched up, say that we have two sets of cells, *G*_*p*_ and *G*_*q*_, respectively. For a pair of cells, *i* ∈ *G*_*p*_ and *j* ∈ *G*_*q*_, found by *k*-nearest neighbour (kNN) search, the cross-attention layers bring about two latent representation vectors, **z**_*i*_ and **z**_*j*_, respectively. Interestingly, the cross-attention layer forces genes in the cell *i* to be associated with the genes in the matched cell *j*, and vice versa.

##### (3) Contrastive learning of common cellular representations

Unlike generative model training, contrastive learning involves few modelling assumptions. Since the latent representation in the bottleneck layer, **z** ∈ ℝ^25^, is somewhat restrictive, we linearly project them onto a large space:

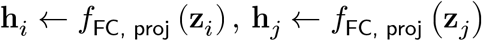

where we use two FC layers–the first and second with 50 units. For each layer, we linearly project the input data and pass it through a series of 1-dimensional batch normalization [36], rectified linear unit (ReLu) activation [37], and dropout regularization [38] with the rate 0.1.

The similarity of two projected vectors, for instance, **h**_*i*_ and **h**_*j*_, can be measured by the following cosine similarity:

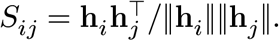

In order to train the model parameters via a self-supervised learning framework [33,39], or noise contrastive estimation (NCE) [32], we need to define positive and negative examples.

Without loss of generality, suppose we pair cells in the sample *p*, hence a cell *i* ∈ *G*_*p*_, with a cell *j* ∈ *G*_*q*_, sampled from the matched sample *q*. We give positive labels, *i* and *j*, to the originally paired cells and find noisy (background) pairs by simply sampling other cell *k* within the same cell group, *G*_*p*_\{*i*}, except for itself. Then, we have similarity scores, *S*_*ij*_, for the positive pairs and the other scores, *S*_*ik*_, for the negative ones. By sampling negative/background cells within the cell group derived from the same donor/patient, we achieve sharp contrasts between the positive and negative pairs.

For each mini-batch, mutual information loss [39] desired by NCE can be bounded by the following:

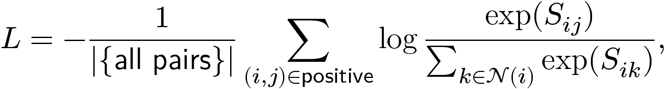

where *N*(*i*) ⊂ *G*_*p*_ is a set of negative examples for a cell *i* found within the same patient group.

#### Learning patient-specific unique representations

Our next question is: *Had we transported a cell into cell-type-less contexts, what would be the patient-specific signatures?* We will address this question by designing a conditional model. Built on a typical variational autoencoder architecture, more precisely, a sufficiently deep encoder and zero-inflated negative binomial (ZINB) decoder proposed by scvi [12], we include all the sources of gene expression variation. The technical details of ZINB models can be found in the original and related papers [13].

#### Model definition

Letting **y**_*i*_ be a latent representation vector, we can design an autoencoder model with the following information flow:

- Encoder: **x**_*i*_ → **y**_*i*_
- Decoder: 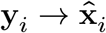

where the reconstructed signals 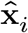 follow ZINB.

Following the idea of the fair autoencoder model [19], we can introduce the latent features **z**_*i*_, each of which can be evaluated (without gradient) by a forward pass of a gene expression vector **x**_*i*_ from the bottleneck layer of the previous network (Fig. 1C).

- Encoder: **x**_*i*_ → **y**_*i*_
- Decoder: 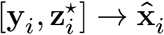

Now, our goal boils down to training a ZINB autoencoder model, while conditioning on the “observed” 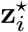 commonality features, so that the new latent representation vector, **y**, can pick up new signatures independent of common cell type signals. Once we have this conditional decoder model, we can predict patient-specific gene expression patterns in a counterfactual manner. For the estimation of cell-type-free, thus patient-specific, perhaps environmental, factors, we can simply switch off **z**, such as Decoder(**y**_*i*_, do(**z**_*i*_ = **0**)).

#### Multi-task learning to delineate the sources of information

In addition to the generative loss of the ZINB model, say *L*^generative^, we enforce strong conditional independence between the two types of latent variables *Y* and *Z* by adding two additional types of loss functions.

All pairwise cosine similarity within a minibatch:

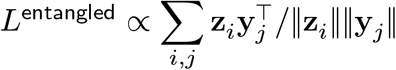

Donor/patient label prediction (cross entropy):

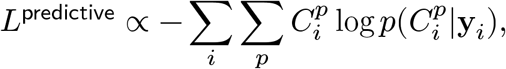

where 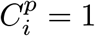 if and only if a cell *i* belongs to a patient *p*, i.e., *C*_p_, otherwise 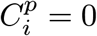.

We may consider putting different weights on different types of losses. Here, we simply equally weighted, constituting the following total loss function:

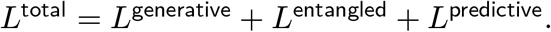

For each loss, we take into account of different number of data points by taking the average using `torch.mean` function.

## Results

### PICASA disentangle sample-specific and sample-invariant effects

As a proof of concept, we first tested PICASA on a well-known batch correction problem, where single-cell expressions of similar pancreatic tissue samples were surveyed by multiple sequencing platforms and technologies. Here, PICASA learnt robust latent representations pertinent to known cell types and states in batch and platform-independent manners (Fig 2A-C). We observed that cells are influenced by both known cell types and batch membership, resulting in many small clusters consisting of cells of the same cell type stemming from one batch (Fig. 2A). We found similar results with ScVi [12]. PICASA clearly separates out two types of latent representations– the sample-specific (Fig. 2B) and commonality (thus cell types) latent space (Fig. 2C). We not only see that cells in the commonality space are located in proximity with other cells from the same cell types (Fig. 2C), while staying away from the different cell types, but we also find that the sample-specific latent features encourage the cells of the same types and batches are grouped together (Fig. 2B).

As another proof of concept, we trained the model on the breast cancer dataset [28]. An autoencoder model trained without patient information generally results in cell clusters of homogeneous cell types and of a single patient (Fig. 2D). Both cell-type variation and patient heterogeneity seem equally dominant, although cell types are generally considered a stronger binding force in a typical batch correction algorithm [5,7]. PICASA, however, resulted in strong cell-type-specific clustering patterns in the UMAP space (Fig. 2E, top), mixing cells of multiple patients together within each cell-type cluster/neighbour (Fig. 2E, bottom). On the other hand, the unique and patient-specific model of PICASA induces tight clustering of malignant cells of the same cancer patient (Fig. 2F, top) and the same subtype (Fig. 2F, bottom). However, interestingly, non-malignant cells (tumour microenvironment) are less patient-specific, forming larger clusters including cells across multiple patients.

Further, we extensively evaluated the quality of latent variables induced by multiple stages of our PICASA framework with seven other state-of-the-art methods, both qualitatively (Fig. S1-7) and quantitatively using five different evaluation metrics (Fig. S8-14) (Supplementary S2). Taken together, benchmark studies suggest that the two latent representations learned by PICASA are empirically identifiable and statistically independent. PICASA is competitive with popular data integration methods while uniquely providing decomposition of shared-residual effects and enabling a more comprehensive characterization of cancer heterogeneity.

### PICASA captures therapy-induced changes in high-grade serous ovarian cancer (HGSOC)

Next, we applied PICASA to ovarian cancer data [29] with a more sophisticated experimental design. This study traced eleven patients between pre- and post-neoadjuvant chemotherapy (NACT). While not explicitly handling patient heterogeneity, autoencoder model fitting resulted in fragmented clustering patterns. Cells generally form well-separated groups by the major cell type categories (Fig. 3A, left), but we also noticed granular structures primarily determined by the patient-of-origin labels (Fig. 3A, right), confirming a large degree of patient heterogeneity, even for non-malignant cells. In the space shared across multiple patient samples, however, we found cells annotated as the same cell type form bigger clusters, one cluster for each cell type, and we also note that relatively minor cell types generally form independent groups (Fig. 3B, left). No batch- or sample-specific clusters are found on the same latent space (Fig. 3B, right, coloured by the patient labels), contrary to the results of the previous autoencoder model (Fig. 3A, right). Next, we examined the patterns of the attention layer within each cell type (Fig. 3C), ascertaining the most frequent gene-gene interactions captured during the model training (Fig. 1) (Supplementary S6). Further, we projected cancer (Fig. 3D) and non-malignant cells (Fig. 3E) separately. The visual inspection of UMAP embedding suggests malignant cells generally form clusters in a highly patient-specific manner, whereas we find non-malignant cells tend to mingle across different patients. Interestingly, for the malignant cells (Fig. 3D, right), we can spot markedly distinctive cell groups within EOC733, EOC87, and EOC372, according to the treatment regimen.

**Fig. 3.**
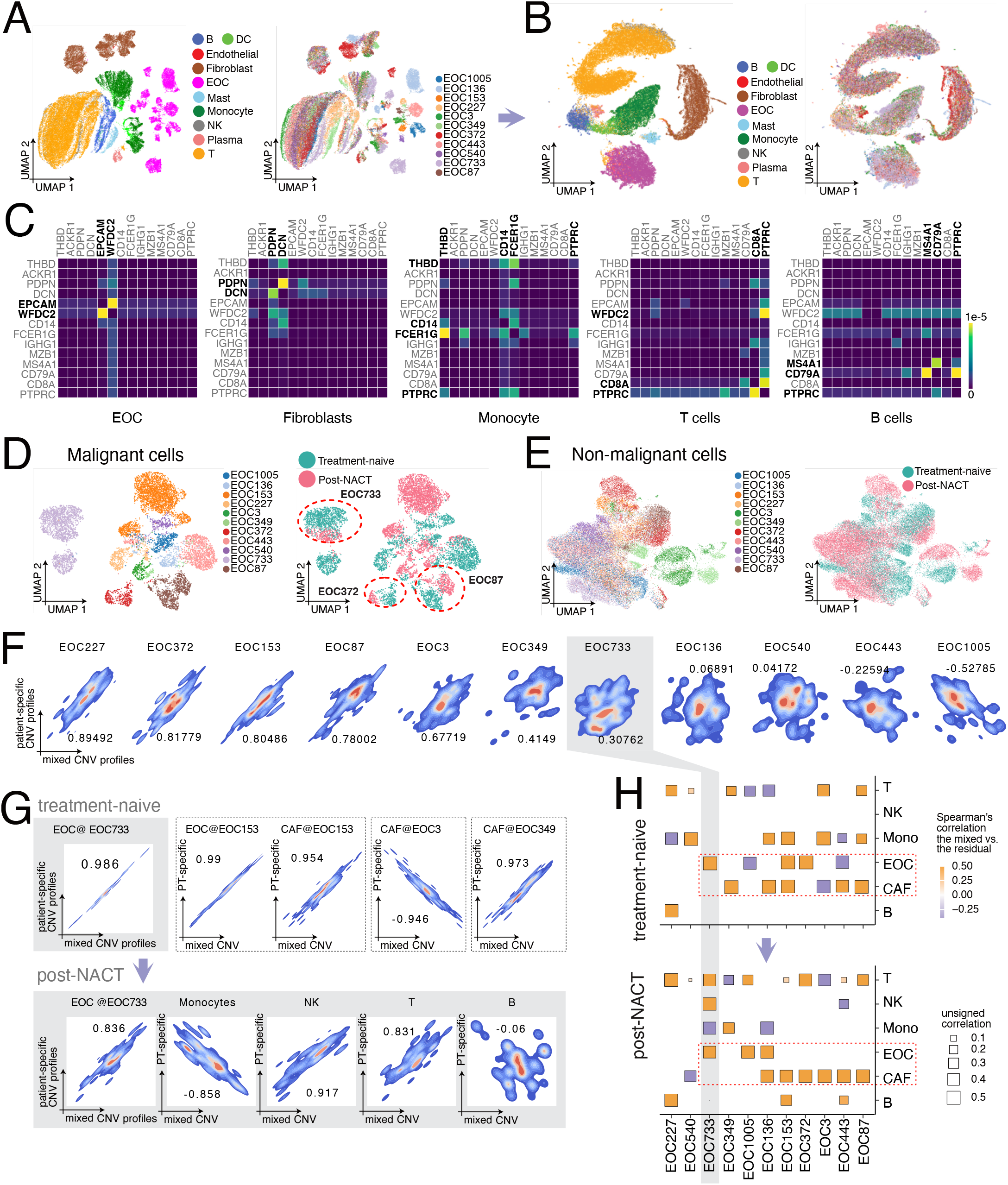
PICASA captures therapy-induced changes in high-grade serous ovarian cancer data. PICASA captures shared cell type and treatment-based patient-specific effects in ovarian cancer. UMAP of **(A)** mixed and decomposed **(B)** common and **(C)** unique representations. **(C)** Heatmap of gene interaction scores of known marker genes for different cell types in the ovarian cancer dataset learned by the cross-attention module. The colours represent row-wise softmax values in the attention matrix of the commonality model. UMAP of **(D)** cancer and **(E)** non-cancer cells from unique representation space learned by the patient-specific network (left panel is patient label and right panel is treatment label). **(F)** Patient-wise contour plots of CNV profiles generated using `copyKAT` from patient-specific and mixed representations of ovarian cancer data. **(G)** Cell type-wise contour plots of CNV profiles for selected patients generated using `copyKAT` from patient-specific and mixed representations. **(H)** Heatmap of correlation between CNV profiles from patient-specific and mixed representations for different cell types at treatment-naive and post-NACT stages.

We then investigated how these patient-specific signals had been generated in light of somatic mutations. Here, we first demonstrate that patient-specific cell signatures are more correlated with the state-of-the-art genomic copy number variants (CNV) profiles using CopyKAT [40], which consistently outperformed in the recent benchmark study [41] (Supplementary S4). Specifically, we first estimated CNV profiles on the mixed (raw) single-cell RNA-seq data and then on the patient-specific transcriptomic signals after eliminating the effect of cell type effects (depicted in Fig. 1C; see Methods for details). Using the two types of cell-level CNV profiles, we first compared them within each patient (Fig. 3F), asking whether the residual signals indeed corresponded to patient-specific genetic effects estimated by CopyKAT or whether there was any possibility that cellular effects (which we removed) were being read as “somatic” copy number changes. For six out of the eleven patients, we found a strong genome-wide correlation between the CNV expressions based on the raw and residual data (R > 0.4); for the other four cases, we found weakly correlated or anti-correlated patterns.

In order to better understand features of residual effects captured by the model, we aggregated the CNV profiles within each cell type and treatment regimen to compare the correlation between the raw and residual data. A majority of copy number changes could be attributed to occur in EOC cells (R = median: 0.89; mean: 0.48 ±0.84), rather than other cell types in the tumour microenvironments, such as the macrophages (R = median: 0.63; mean: 0.18 ±0.83), the T-cells (R = median: 0.52; mean: 0.26 ±0.6), and the B-cells (R = median: 0.65; mean: 0.57 ±0.41) (Fig. 3G). In patient-specific analysis, EOC773 measured substantially strong Spearman’s correlation between the raw and residual CNV values before treatment (R=0.99) and the majority of cell populations were considered the EOC type. After the neoadjuvant chemotherapy (NACT), we still found a strong correlation for the EOC (R=0.84). However, we also found that non-malignant cells report high correlation values within their own cell type, such as the cases of T-cells (R=0.83) and the natural killer (NK) cells (R=0.83). Interestingly, we found an inverse correlation for the macrophages (R=-0.86 and very weak correlation for the B-cells (R=-0.06). For some of the cancer-associated fibroblasts (CAF) samples, we also found strong correlations, such as EOC349 (R=0.97) before treatment, EOC136 (R=0.86), EOC153 (R=0.84), EOC3 (R=0.95), EOC443 (R=0.94), and EOC87 (R=0.95) after treatment. Arguably, CNV results relying on gene expression profiles are not the best way to track cancer clonal status. Nonetheless, it is reassuring to see generally stronger correlations among the malignant cell types.

### PICASA characterizes subtypes of non-small cell lung cancer (NSCLC) cells

Similarly, in the lung cancer studies, PICASA consistently map 72,514 cells derived from 23 NSCLC patients, encompassing both lung adenocarcinoma (LUAD) and lung squamous cell carcinoma (LUSC) [30], and strongly induces distinctive cell type clusters while not showing any patient-specific effects (Fig. 4A). Each cell type’s commonality is characterized by interactions between marker genes (Fig. 4B). Further, we conducted a gene set enrichment analysis of highly variable genes (2,000 genes) within each common cell type cluster against the marker genes in PanglaoDB [42] using rank-based Fast Gene Set Enrichment Analysis (GSEA) [43] implemented in R’s fgsea package. Here, we show that cell-type-specific cross-attention activities also recapitulate known cell-type marker genes (Fig. 4C) (Supplementary S6). High-activity genes in the endothelial cluster are significantly over-represented in endothelial cells (p=0.001), stromal cells (p=0.008), and precursor mesothelial cells (p=0.008). We found that the top genes in the fibroblast cluster enrich mesangial cells (p=0.005) and myofibroblast cells (p=0.006). For the malignant cells, our GSEA results highlight the true cell type of origin, which are epithelial cell types, such as alveolar goblet cells (p<0.001), epithelial cells (p=0.001), and basal cells (p=0.005). We observe similar enrichment patterns for the non-cancer epithelial cells, whose gene activity ranks are associated with goblet cells (p=0.015), epithelial cells (p=0.02), alveolar cells (p=0.03), and basal cells (p=0.04). As for the plasma cells and other immune cells, we found the plasma cell clusters match with plasma (p=0.001)/B cells (p=0.006), the monocyte/macrophage cell cluster with alveolar macrophages (p=0.009), and T-cells with T-cells (p=0.002), cytotoxic T-cells (p=0.002), helper T-cells (p=0.006) and naive T-cells (p=0.04).

**Fig. 4.**
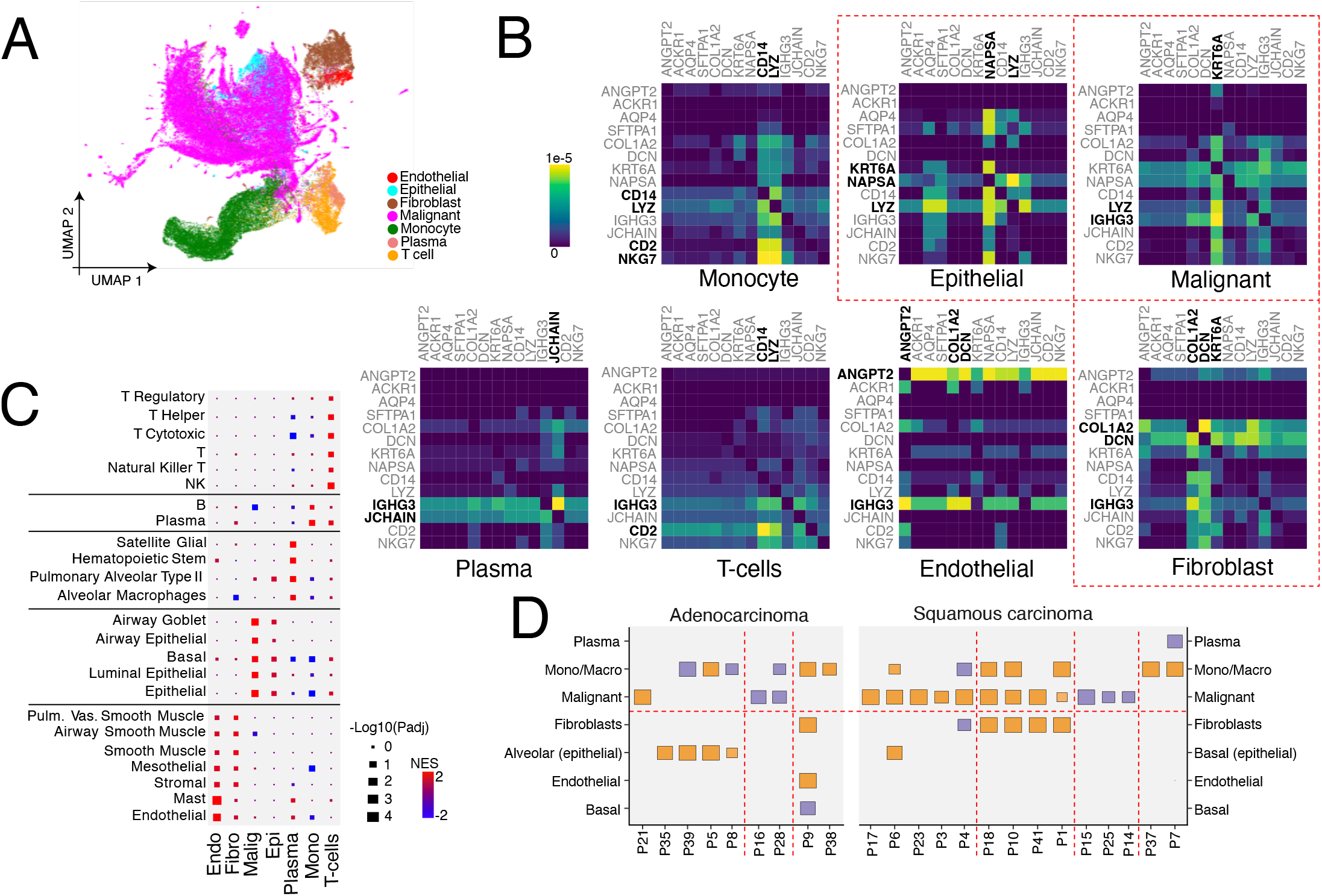
PICASA characterizes subtypes of non-small cell lung cancer. PICASA captures shared cell type and histologically defined cancer subtype-specific effects in lung cancer. **(A)** UMAP of decomposed common representation with cell type label. **(B)** Heatmap of gene interaction scores of known marker genes for different cell types in the lung cancer dataset learned by the cross-attention module. **(C)** Gene set enrichment analysis of lung cancer data based on ranked genes in the cross-attention matrix with the known cell-type marker genes in the PanglaoDB database. **(D)** Heatmap of correlation between CNV profiles from patient-specific and mixed representations for different cell types.

As we did in the ovarian cancer study, we summarized Spearman’s correlation between the two types of CNV profiles estimated on the raw (mixed) and residual (patient-specific) expression, respectively. For the Alveolar cells derived from the four LUAD (adenocarcinoma) patients, we found strong correlations (median: 0.9; mean: 0.81 ±0.26), suggesting that the genetic effects of the cells in these patients had not been “adjusted” by cell type effects (thus, independent). Likewise, we found strong positive correlations for malignant cells derived from LUSC (squamous carcinoma) patients (eight out of thirteen), yielding R=median: 0.89; mean: 0.47 ±0.74. In summary, many of the genetic effects were seen in alveolar cell clusters for LUAD patients, whereas the genetic effects of malignant cells were quite robustly sustained in LUSC patients. However, interestingly, we found negative correlation values for P39 and P8 (adenocarcinoma) for the monocytes/macrophages, respectively, R=−0.93 and −0.56. We also found negative correlations for the malignant cells in P16 and P28, R=−0.79 and −0.69, respectively. Such negative correlations between the raw and residual data highlight that single-cell data alone measured within each patient and cell type may lack a necessary basis to discern transcriptomic signals derived from genetic/somatic effects and developmental/differentiation processes.

## Discussion

We note several limitations of this study. One of our key ideas in neural network architecture builds on a simplified version of a transformer network [35]. For simplicity, we only implemented expression embedding schemes, restricting the 2,000 most highly variable genes/features in modelling gene-gene interaction attention matrices. In many cases, unless our goal is to build a foundation model, a thousand genes often suffice to capture general cell type signatures. However, we can increase the number of features in our cross-attention layers by adopting gene symbol-based embedding and tokenization as in other single-cell foundation models [25,27], where we restrict the number of tokens with different genes for different cells. Moreover, the attention mechanism layer itself can be more efficiently implemented, reducing memory footprint and computation cost [44]. More efficient transformer architectures will enable us to include ten times more features in the model [45].

We intentionally designed our approach to take two steps in getting final patient-specific expression values, later used to make genome-wide CNV calls. In our preliminary analysis, resolving both shared and patient-specific latent representations poses challenges in model identifiability. In fact, without enforcing a strong prior probability, finding latent variables mutually independent of one another is a non-trivial problem [46–48], perhaps an open question. In a sense, estimating the parameters of a complete joint model would necessarily involve somewhat arbitrary and laborious hyperparameter optimization. Instead, joint analysis with genomic data (DLP+) [49] and probabilistic mapping between DNA and RNA samples [50] is expected to pave a new avenue to more principled approaches. Alternatively, we can preprocess to have single-nucleotide variant calls and read depth profiles from RNA-seq data to make sure that different data modalities capture genetic effects.

## Data Availability

All the datasets used in this study are publicly available in Gene Expression Omnibus (GEO) database.

https://www.ncbi.nlm.nih.gov/geo/query/acc.cgi?acc=GSE148071

https://www.ncbi.nlm.nih.gov/geo/query/acc.cgi?acc=GSE165897

https://www.ncbi.nlm.nih.gov/geo/query/acc.cgi?acc=GSE176078

