## Supplementary for "PICASA: Decomposing patient heterogeneity of single-cell cancer data by cross-attention neural networks"

#### Contents

**S1 Simulation datasets preprocessing** **2**

**S2 Benchmark analyses using simulated and real-world datasets** **3**

**S3 Construction of Matched Cell Pairs** **22**

**S4 Copy Number Variation (CNV) analysis** **22**

**S5 Computational architecture and scalability** **22**

**S6 Cross-attention module reveals gene interactions in normal and cancer datasets** **27**

**Reference** **40**

### S1 Simulation datasets preprocessing

We simulated single-cell data sets in the following three different scenarios.

- Simulation #1 (small-scale, balanced): For the total of 8,700 cells, we partitioned them into three equal-sized batches (2,900 cells per batch) and independently into five different cell type groups with an equal probability (20%). We simulated 1,000 genes per cell using the default forward simulator implemented in `Splatter` library [1].
- Simulation #2 (large-scale, balanced): We also considered a different type of simulation scheme proposed in the atlas-level batch integration benchmark study [2]. For the total of 19,318 cells, we first defined equally-sized (technical) batches and independently grouped them into four cell-type groups. The batch membership was provided to unsupervised learning algorithms, but the cell-type labels were held out for benchmark evaluation. For simplicity, we focused on 2,000 genes that are highly expressed and thus consistently available for all the cells.
- Simulation #3 (large-scale, unbalanced): We simulated more challenging benchmark data using the same simulation scheme [2] as above but with skewed cell distributions across batches and cell-type groups. We used 2,000 highly expressed (or variable) genes. For six technical batches, we broke down the total of 12,097 cells as follows: ($\approx$ 2,900, $\approx$ 2,500, $\approx$ 2,000, $\approx$ 1,900, $\approx$ 1,700, $\approx$ 1,000). For the cell-type allocation, we independently assigned the cells to one of the seven cell types with the following probability: (0.3, 0.23, 0.15, 0.12, 0.1, 0.05, 0.01).

The simulated data sets #2 and #3 had been generated and made accessible through Figshare platform.

### S2 Benchmark analyses using simulated and real-world datasets

To evaluate the integration performance of PICASA, we analyzed simulated, normal, and cancer datasets using the seven state-of-the-art integration methods and compared the results with PICASA method. We used `scanpy` package [3] as the base tool for preprocessing and highly variable gene selection. Next, we used the obtained latent space representation with the same number of dimensions using `scanpy` API or an independent environment installation according to the respective tool documentation. Finally, to generate the neighbour graph and UMAP visualization, we used `scanpy` for all methods, including PICASA.

We compared the quality of latent variables induced by multiple stages of our PICASA framework with other methods both qualitatively (Figures S1, S2, S3, S4, S5, S6, S7) and quantitatively (Figures S8, S9, S10, S11, S12, S13, S14).

The methods used for comparison include the following-

**PCA** PCA-based data integration implemented in Scanpy [4]

**Combat** (version 3.4.0) factorization-based batch adjustment method [5]

**BBKNN** (version 1.6.0): batch-balancing K-nearest neighbour [6]

**Harmony** (version 0.0.10): cluster-based batch adjustment [7]

**Scanorama** (version 1.7.4): mutual nearest neighbour method [8]

**Liger** (version 0.2.3): non-negative matrix factorization method [9,10]

**scVI** (version 1.2.1): variational autoencoder model with batch labels [11]

**PICASA-C** (this work): latent commonality space

**PICASA-U** (this work): residual, sample-unique space

**PICASA-UC** (this work): the commonality and residual states combined

We confirmed that the performance of data integration by PICASA model is qualitatively and quantitatively competitive with other existing state-of-the-art integration methods. We projected the latent states estimated by different methods in two-dimensional UMAP space and coloured each cell according to cell types (lower panel) and samples (upper panel) (Figures S1, S2, S3, S4). BBKNN, Liger, and PICASA-C consistently formed a single (nearly perfect) cluster for each cell type group across different datasets. The other methods, such as Scanorama, scVI, and COMBAT, generally place one cell type in one cluster but often result in disjoint and scattered, tiny clusters that are unable to reconcile the differences between samples. Clusters generated by PICASA-C are tightly clustered within each cell type while mixing multiple samples. On the other hand, PICASA-U works in the opposite way, mixing multiple cell types while separating samples and forming sample-specific clusters. For cancer datasets, Harmony, scVI, and PICASA-C methods consistently performed better, and the integrated UMAP representations did not show any patient-specific clusters (Figures S5, S6, S7).

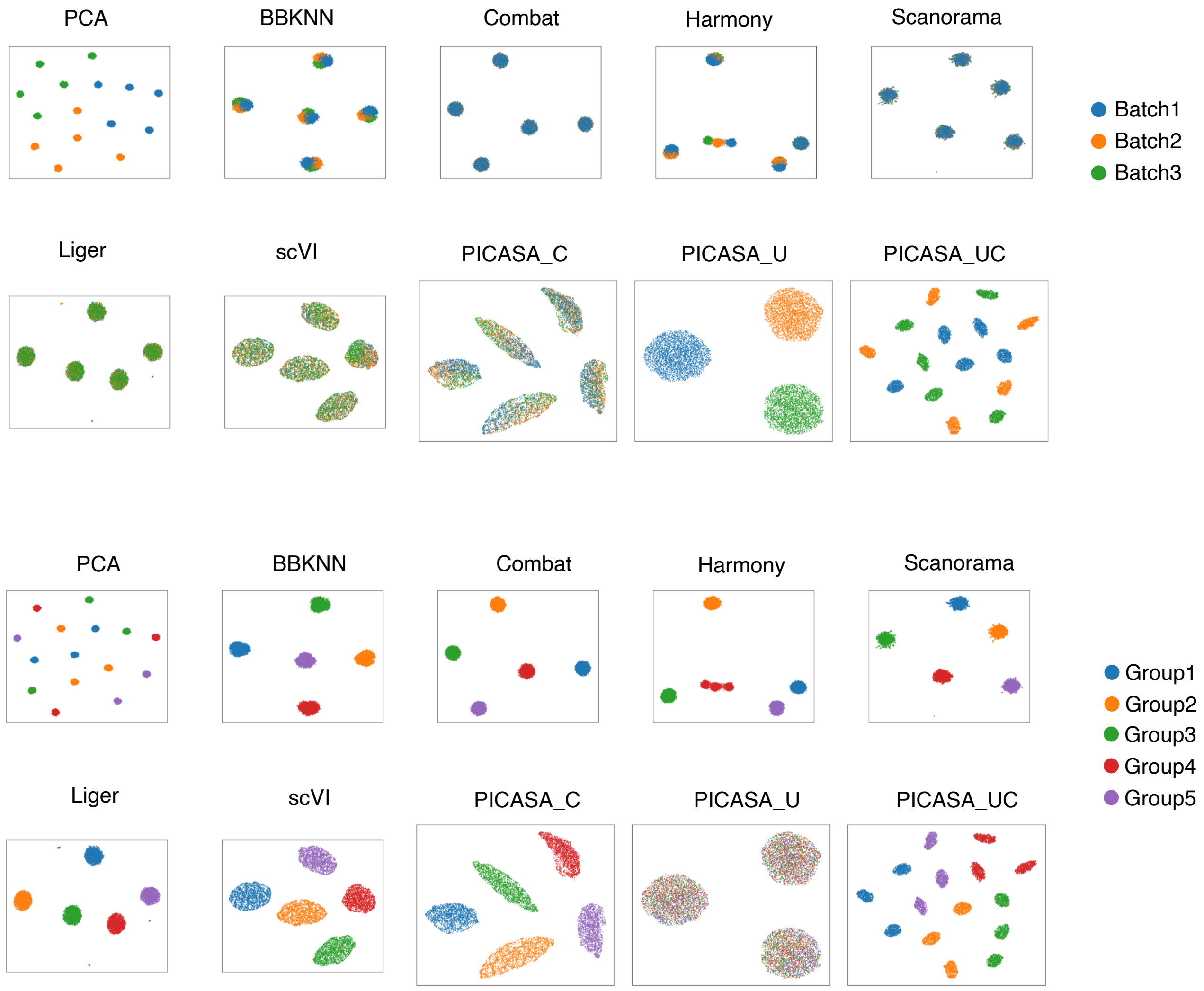

**Fig. S1**: Comparative UMAP representation of PICASA capturing shared cell type effects and integrating cells across samples in Simulation-1 (small-scale, balanced) dataset.

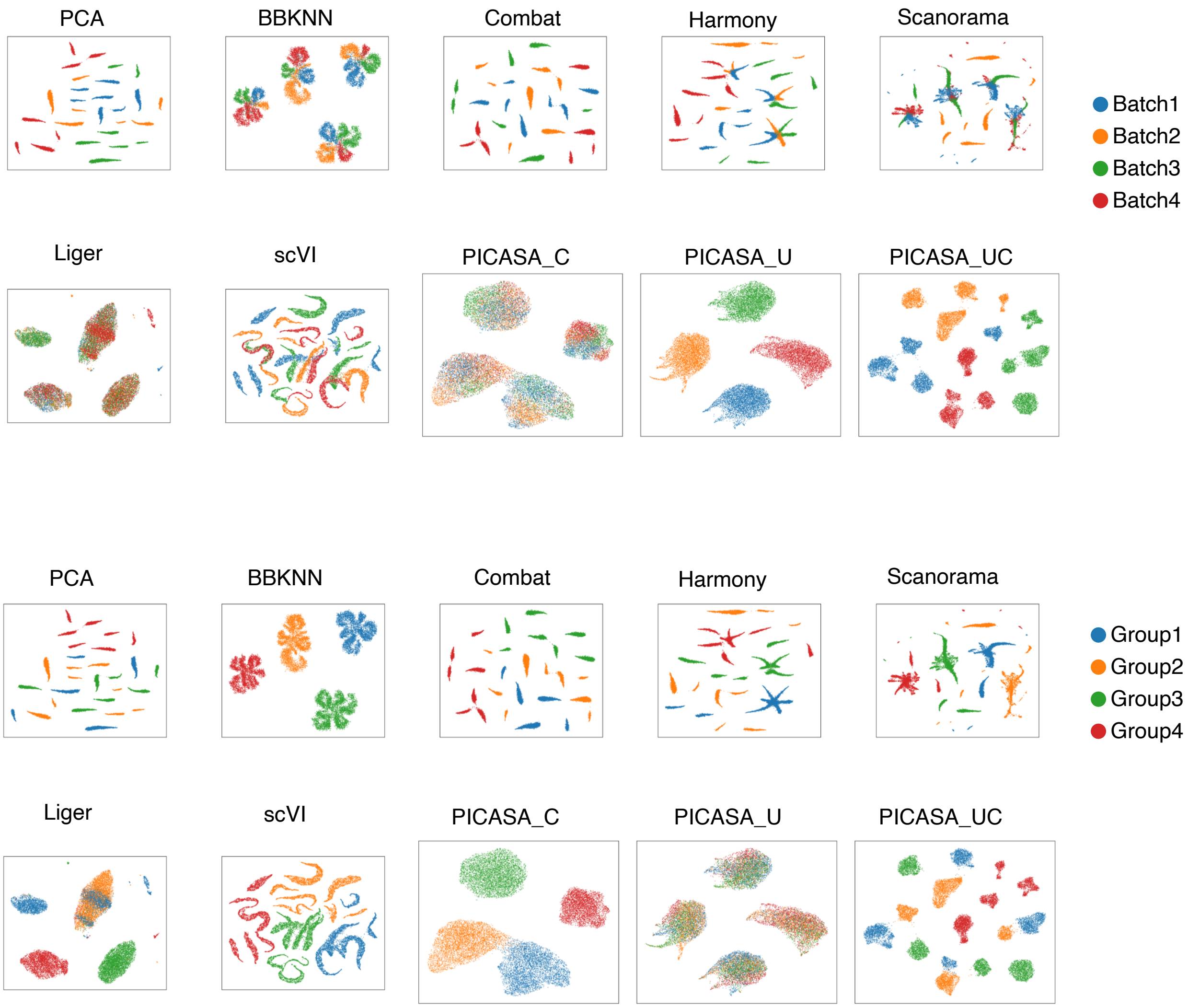

**Fig. S2**: Comparative UMAP representation of PICASA capturing shared cell type effects and integrating cells across samples in Simulation-2 (large-scale, balanced) dataset.

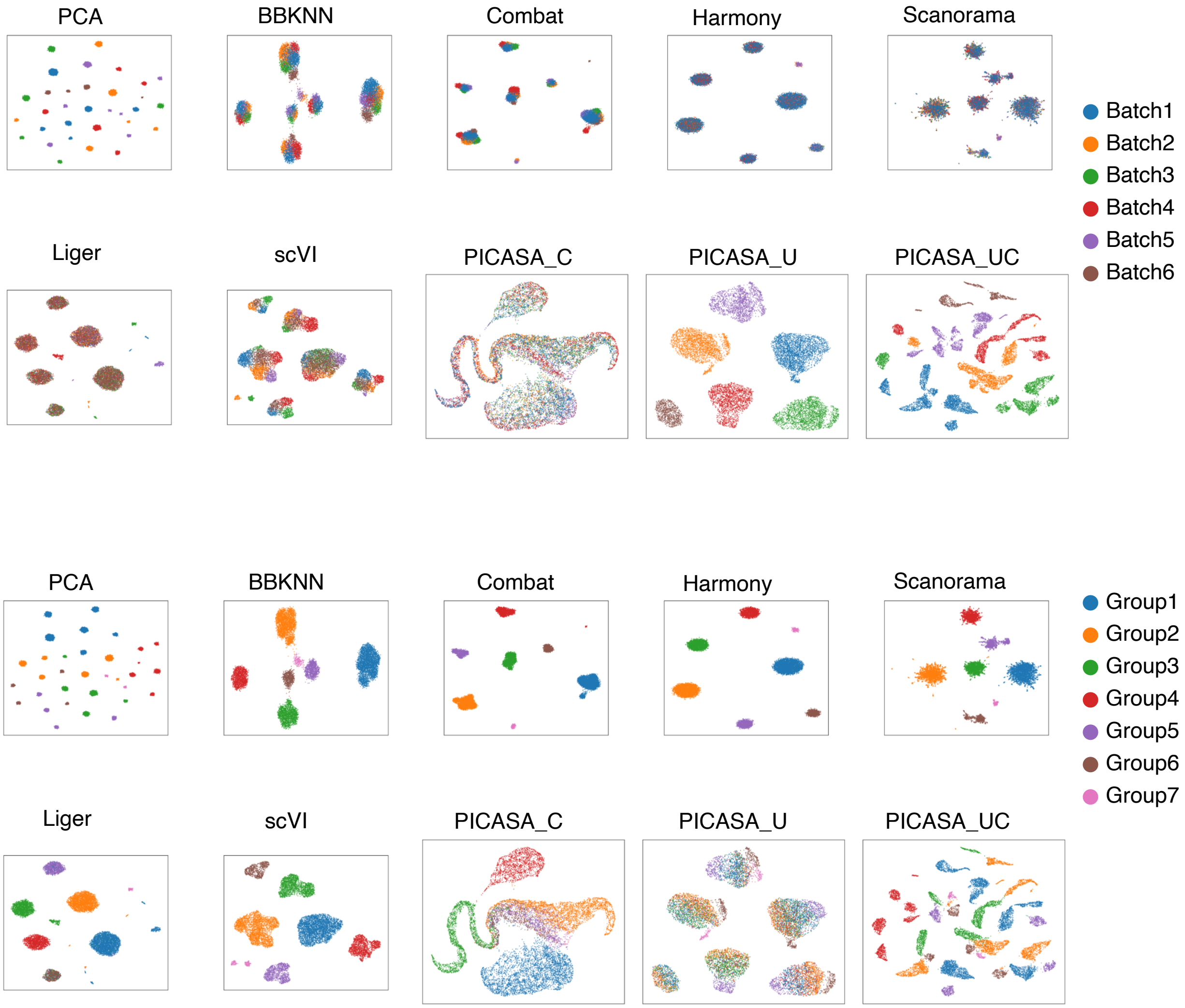

**Fig. S3**: Comparative UMAP representation of PICASA capturing shared cell type effects and integrating cells across samples in Simulation-3 (large-scale, unbalanced) dataset.

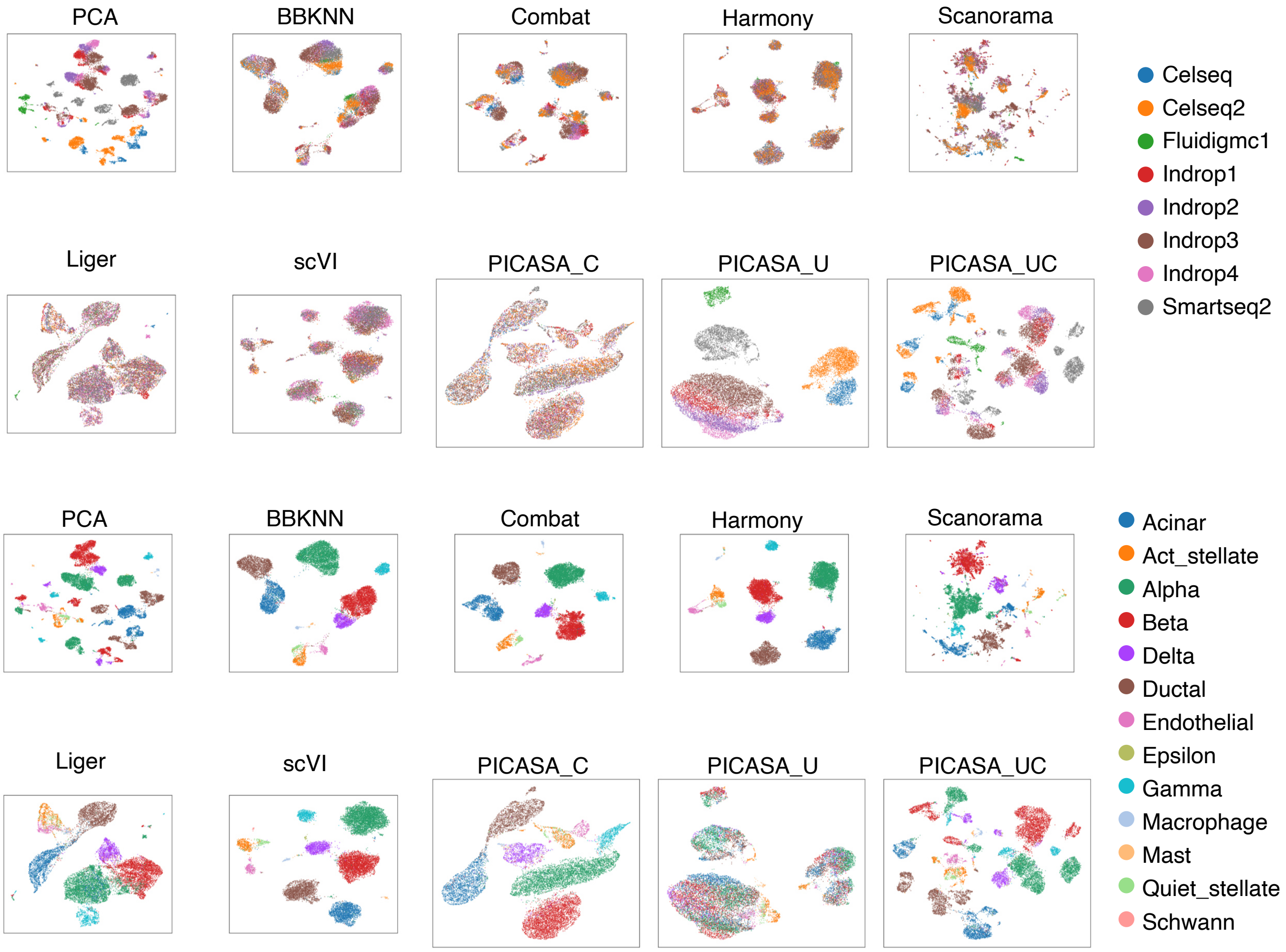

**Fig. S4**: Comparative UMAP representation of PICASA capturing shared cell type effects and integrating cells across samples in normal pancreas dataset.

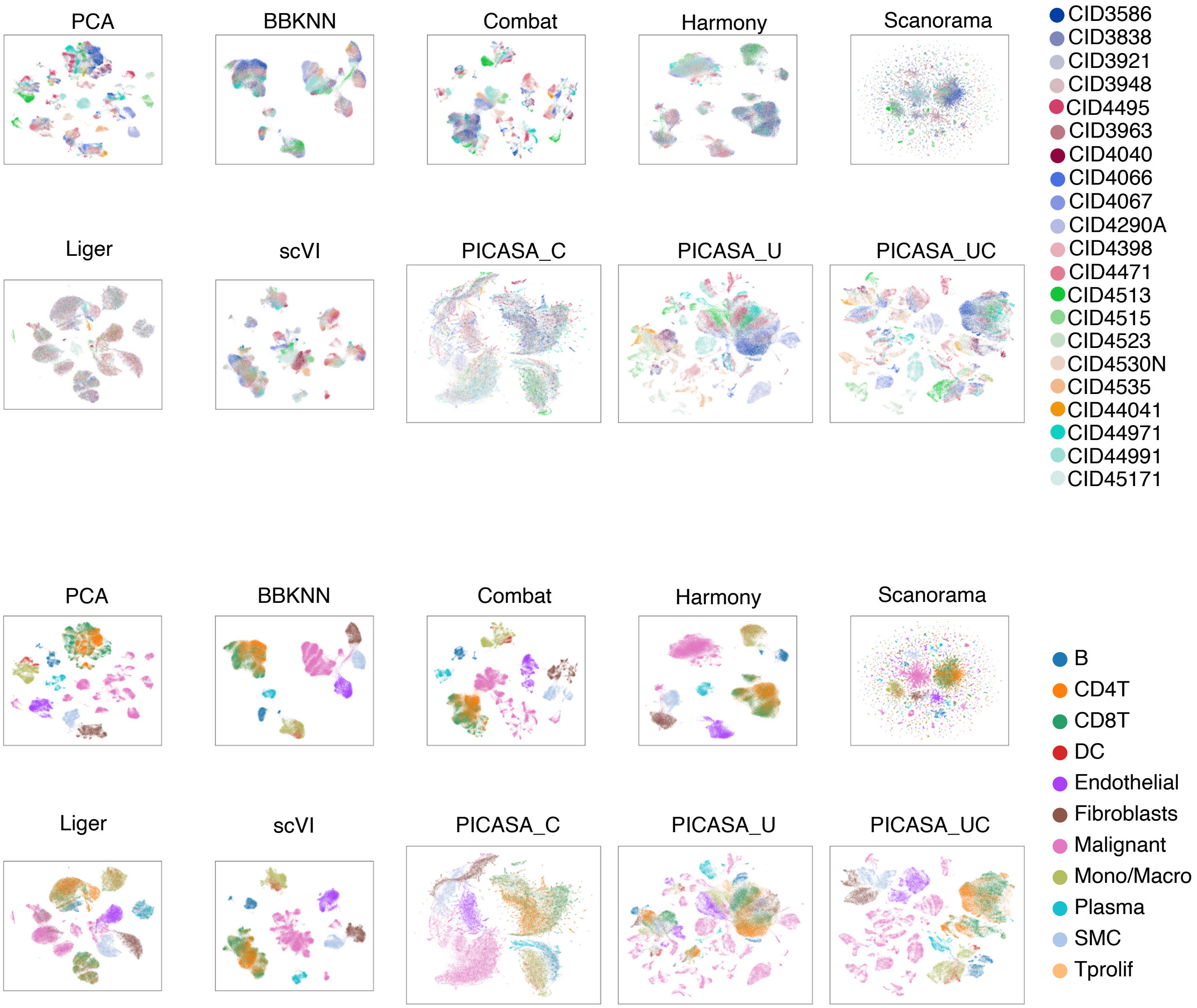

**Fig. S5**: Comparative UMAP representation of PICASA capturing shared cell type effects and integrating cells across samples in breast cancer dataset.

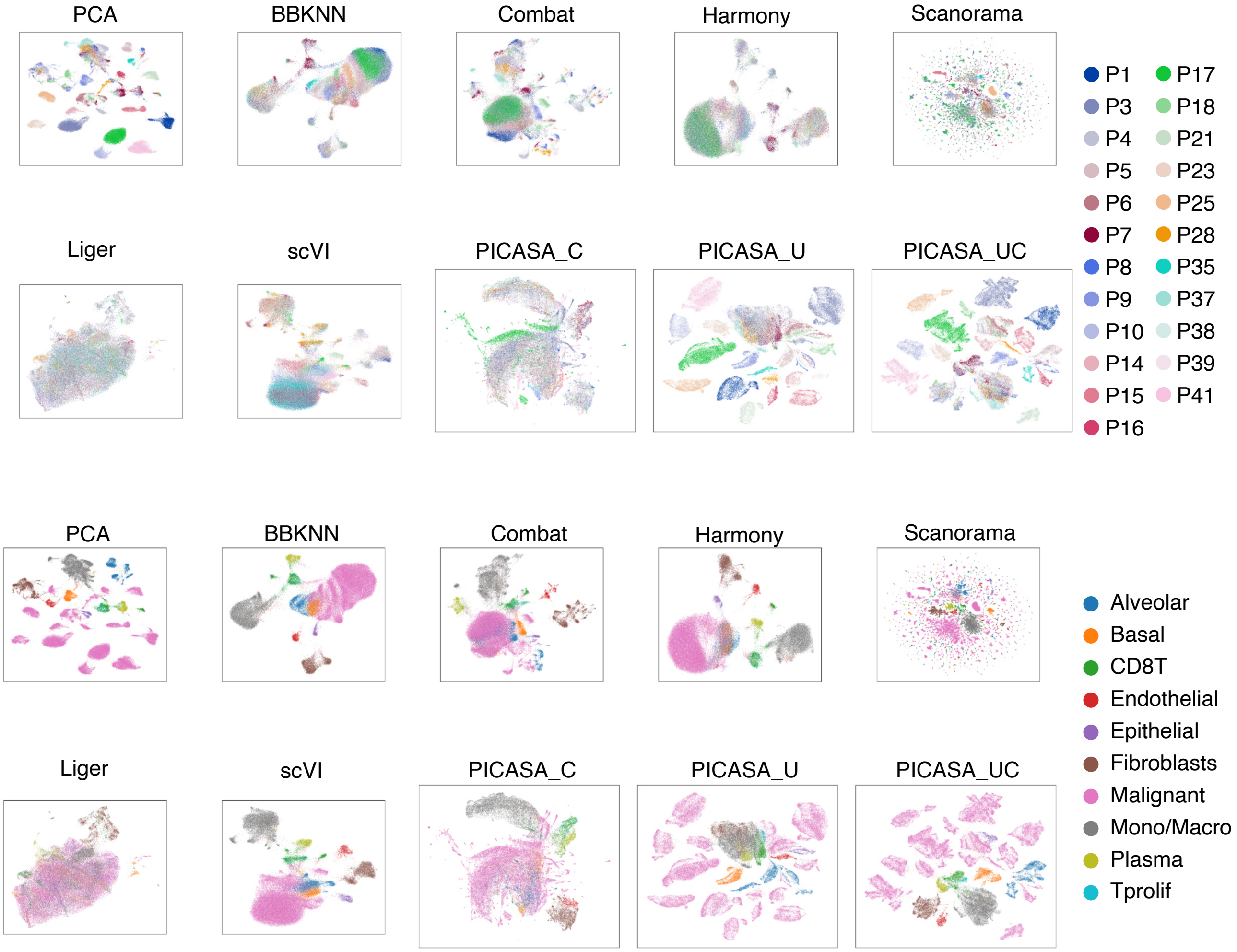

**Fig. S6**: Comparative UMAP representation of PICASA capturing shared cell type effects and integrating cells across samples in lung cancer dataset.

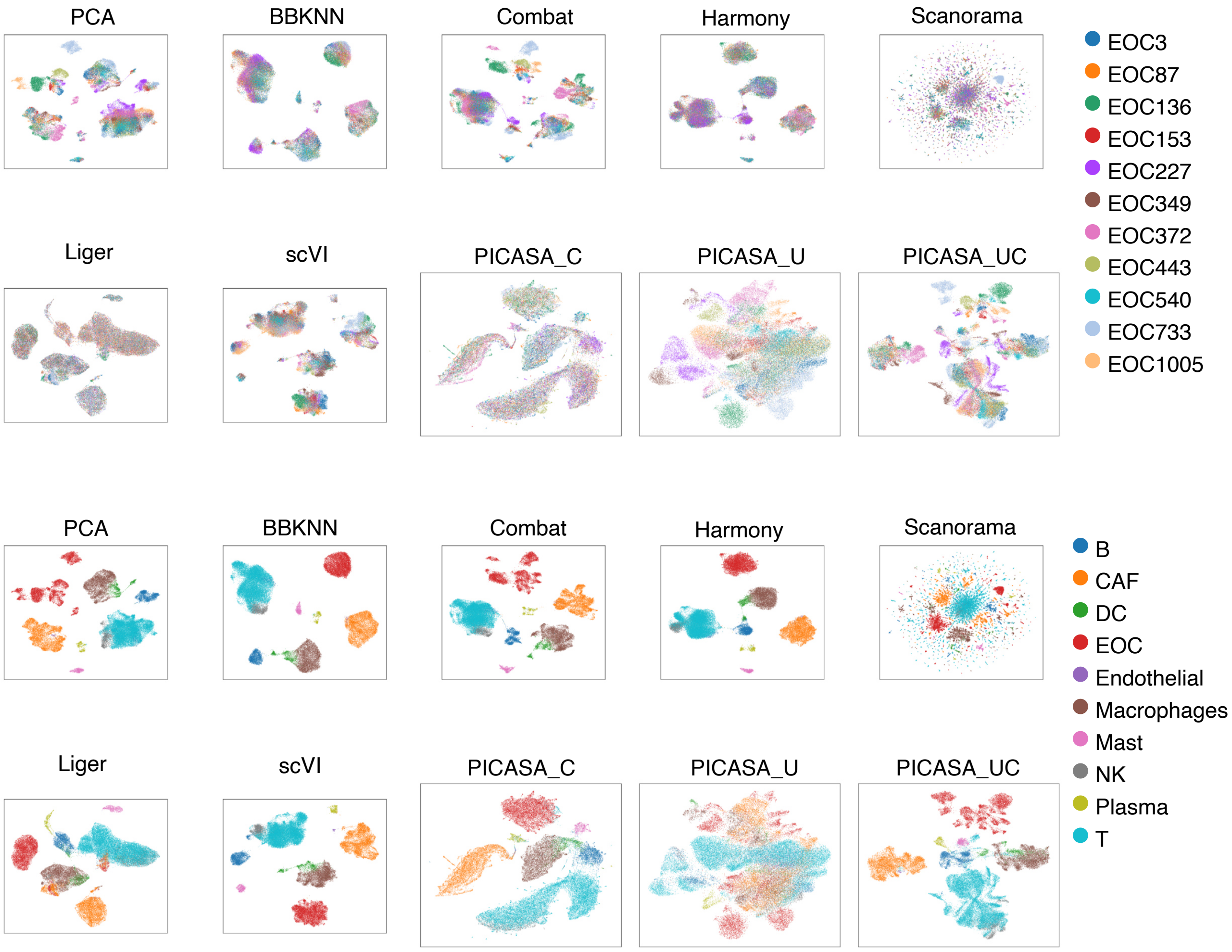

**Fig. S7**: Comparative UMAP representation of PICASA capturing shared cell type effects and integrating cells across samples in ovarian cancer dataset.

We then quantified how well PICASA and other methods perform in data integration tasks using five different evaluation metrics- LISI (Local Inverse Simpson's Index) [12], Silhouette score [13], Connected Components score [14], Adjusted Rand Index (ARI) [15], and Normalized Mutual Information (NMI) [16] (Figures S8, S9, S10, S11). The LISI (Local Inverse Simpson's Index) scores [12], namely, $1/\sum_{i=1}^{n} p_i$ where $p_i$ is the probability of cell $i$'s neighbours belonging to the same cell type or batch label. We used the implementation from *compute-lisi* function in the `harmonypy` (version 0.0.10) package and described in the original `harmony` integration method paper [12]. The LISI evaluation selects several neighbours based on a predefined perplexity (default 30) and calculates a score to test how well cells from different conditions are mixed. To compare the methods, we calculated the LISI score for each cell and calculated an average score across all the cells in the data. A low LISI value means a homogeneous mixture of neighbouring cells with the minimum value of 1; a high LISI implies the opposite, heterogeneity within the neighbours. To evaluate data integration performance across different batches or patients, we calculated the LISI score based on batch or patient label. Our expectation for PICASA-C is to have low LISI values with respect to cell type labels and high LISI regarding batch/sample labels. For PICASA-U, we expect the opposite- high cell type LISI and low batch LISI. All the methods we considered here show strong performance in terms of grouping cells into clusters of a homogeneous cell type, notably for the batch LISI, Liger and PICASA-C consistently performed better than other methods (top panel in Figures S8, S9, S10, S11). For simulation #1, COMBAT achieved the highest batch LISI score (mean: $2.62 \pm 0.24$), outperforming PICASA-C (mean: $2.27 \pm 0.43$). For simulation #2, Liger yielded the highest batch LISI score (mean: $2.69 \pm 0.66$), compared to PICASA-C (mean: $2.16 \pm 0.72$). Similarly, for simulation #3, Liger again obtained the highest batch LISI score (mean: $3.60 \pm 0.84$), while PICASA-C scored $3.02 \pm 0.83$. Across all three simulations, cell LISI scores were consistently close to 1 for all methods, indicating well-mixed cell types. For the normal pancreas dataset, Liger achieved the highest batch LISI score (mean: $3.67 \pm 1.16$), followed by PICASA-C (mean: $3.21 \pm 1.14$), with cell LISI scores again close to 1 across methods.

Next, we evaluated integration quality using Silhouette scores [13] based on both cell type and batch labels. The Silhouette score measures how similar a cell is to others within the same group compared to those in different groups, with values close to 1 indicating well-defined clustering and values near 0 or negative suggesting poor mixing. We used the *silhouette_samples* function in the `scib_metrics` (version 0.5.1) package and described in the original single-cell integration benchmarking paper [17]. To allow consistent interpretation across both label types, we scaled values between 0 and 1 as follows. For cell type labels, we applied a (score+1)/2 transformation, where higher values indicate better separation of cell types. For batch labels, we scaled the raw scores with a 1−absolute(score) transformation such that higher values reflect better batch mixing, i.e., cells from different batches are well-integrated. Here, compared to other methods, along with BBKNN, COMBAT, and Liger, PICASA-C showed strong performance in terms of integrating similar cells into distinct cell type-specific clusters (middle panel in Figures S8, S9, S10, S11). For Simulation #1, COMBAT achieved the highest batch score (mean: $0.99 \pm 0.01$) compared to PICASA-C (mean: $0.88 \pm 0.04$), and BBKNN showed the best cell type score (mean: $0.95 \pm 0.03$) compared to PICASA-C (mean: $0.86 \pm 0.07$). For Simulation #2, the highest batch score was achieved by PICASA-C (mean: $0.90 \pm 0.02$), followed by Liger (mean: $0.88 \pm 0.06$), and the highest cell type score is BBKNN (mean: $0.91 \pm 0.02$) compared to PICASA-C (mean: $0.73 \pm 0.01$). For Simulation #3, COMBAT attained the highest batch score (mean: $0.98 \pm 0.01$) compared to PICASA-C (mean: $0.93 \pm 0.08$), and BBKNN had the highest cell type score (mean: $0.78 \pm 0.20$) relative to PICASA-C (mean: $0.60 \pm 0.09$). Similarly, for the normal pancreas dataset,

COMBAT again showed the best batch correction (mean: 0.90 ±0.04) compared to PICASA-C (mean: 0.79 ±0.06), while BBKNN achieved the highest cell type score (mean: 0.79 ±0.06) compared to PICASA-C (mean: 0.68 ±0.56).

To further evaluate the integration performance of PICASA-C, we calculated graph connectivity using Connected Components analysis on the k-nearest neighbour (kNN) graph [14,18]. We used the *connected_components* function in the `scipy` (version 0.5.1) package and described in the original single-cell integration benchmarking paper [17,19]. First, we generate a global kNN graph based on the latent space representation of all cells and evaluate how well labels are connected within the graph. A score close to 1 indicates that most cells of the same type are connected, while lower values suggest cells from the same cell type are disconnected. Here, PICASA-C consistently performed among the top methods in integrating similar cells within a connected component of a graph (Simulation #1 mean: 0.99 ±0.02, Simulation #2 mean: 0.99 ±0.03, Simulation #3 mean: 0.91 ±0.14, and normal pancreas mean: 0.89 ±0.12) (lower left panel in Figures S8, S9, S10, S11).

Additionally, to evaluate the clustering results in the integrated space and known cell type labels, we calculated the Adjusted Rand Index (ARI) [15] and Normalized Mutual Information (NMI) [16]. We used the *nmi_ari_cluster_labels_leiden* function in the `scib_metrics` (version 0.5.1) package and described in the original single-cell integration benchmarking paper [17]. ARI measures the agreement between the clustering and true labels while adjusting for chance, whereas NMI quantifies the amount of shared information between them. Scores close to 1 indicate strong concordance with biological identity. For PICASA-C, we expect high ARI and NMI scores, reflecting the preservation of cell-type structure. For PICASA-U, we expect low scores, indicating disrupted biological grouping. Here, COMBAT, Harmony, scVI, and PICASA-C showed superior performance across all datasets, except for the unbalanced simulated dataset, where PICASA-C scores were average possibly due to lack of contrastive signals from highly unbalanced/low proportioned simulated data group labels (Simulation #1 ARI: 0.99, NMI: 0.99, Simulation #2 ARI: 0.98, NMI: 0.97, and Simulation #3 ARI: 0.50, NMI: 0.66) (lower right panel in Figures S8, S9, S10, S11). While for real data with an unbalanced class represented by a normal pancreas dataset, the performance of PICASA-C (ARI: 0.91, NMI: 0.86) was comparable to other top-performing methods. For cancer datasets, PICASA-C consistently performed similarly to other top-performing methods in all of the metrics evaluated. However, cluster evaluation for lung cancer data using ARI and NMI for PICASA-C showed low scores mainly due to failure to cluster subtypes of epithelial cells, such as basal and alveolar cells, into distinct clusters (Figures S12, S13, S14). Taken together, the above benchmark experiments suggest that the two types of latent variables that PICASA generates are empirically identifiable and statistically independent from each other.

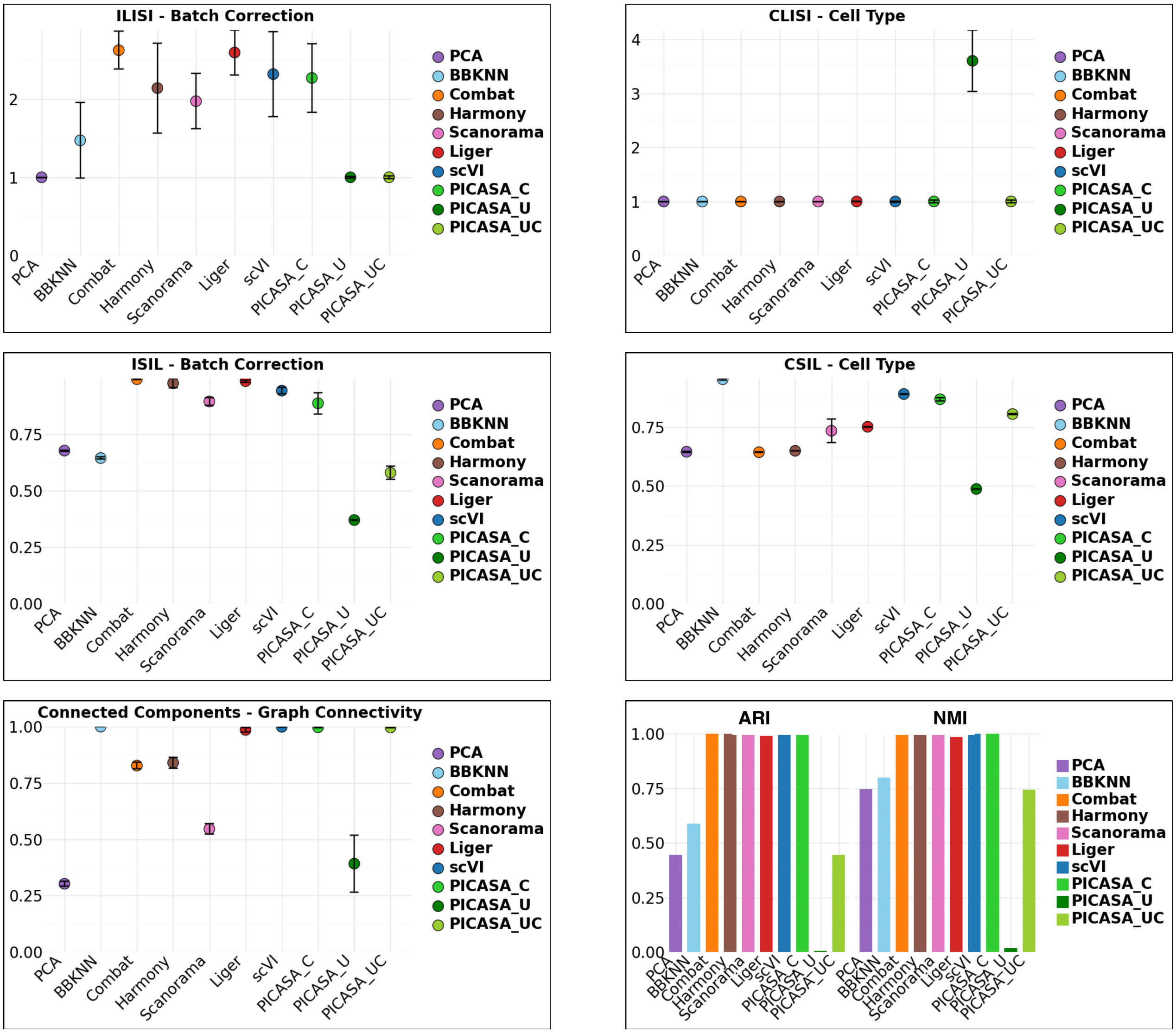

**Fig. S8**: Comparative analysis of PICASA integrating cells across patients in Simulation-1 (small-scale, balanced) dataset. The upper panel plot shows batch label Local Inverse Simpson's Index (LISI) (higher value suggests better mixing of cells across samples) and cell type label Local Inverse Simpson's Index (LISI) (lower value suggests better mixing of cells from the same cell type label). The middle panel plot shows batch label scaled Silhouette score (higher value suggests better mixing of cells across samples) and cell type label scaled Silhouette score (higher value suggests better mixing of cells from the same cell type label). The lower panel plot shows cell type label graph connected component analysis (higher value suggests better mixing of cells across samples), and Adjusted Rand Index (ARI) and Normalized Mutual Information (NMI) score (higher value suggests better mixing of cells from the same cell type label).

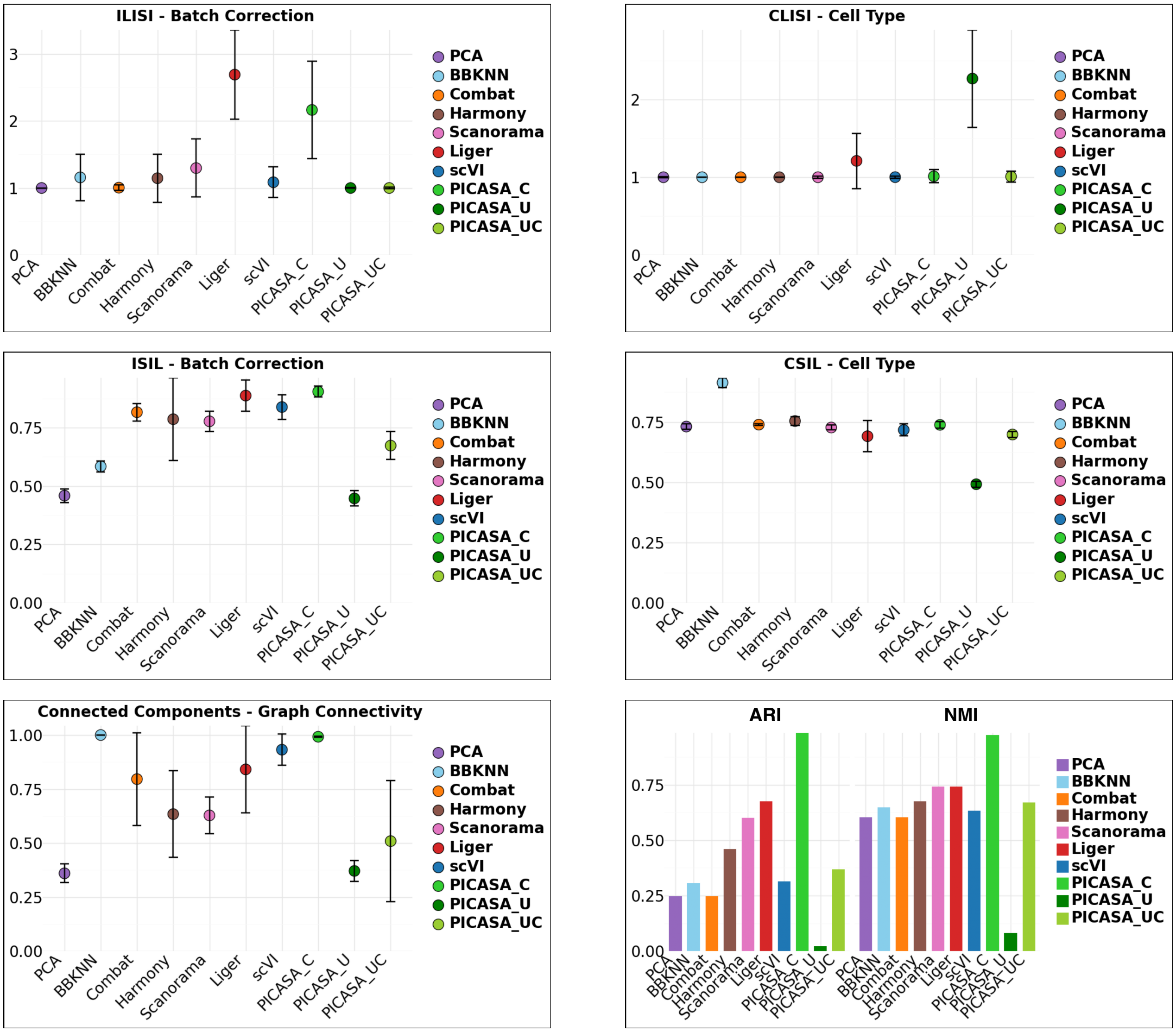

**Fig. S9**: Comparative analysis of PICASA integrating cells across patients in Simulation-2 (large-scale, balanced) dataset. The upper panel plot shows batch label Local Inverse Simpson's Index (LISI) (higher value suggests better mixing of cells across samples) and cell type label Local Inverse Simpson's Index (LISI) (lower value suggests better mixing of cells from the same cell type label). The middle panel plot shows batch label scaled Silhouette score (higher value suggests better mixing of cells across samples) and cell type label scaled Silhouette score (higher value suggests better mixing of cells from the same cell type label). The lower panel plot shows cell type label graph connected component analysis (higher value suggests better mixing of cells across samples), and Adjusted Rand Index (ARI) and Normalized Mutual Information (NMI) score (higher value suggests better mixing of cells from the same cell type label).

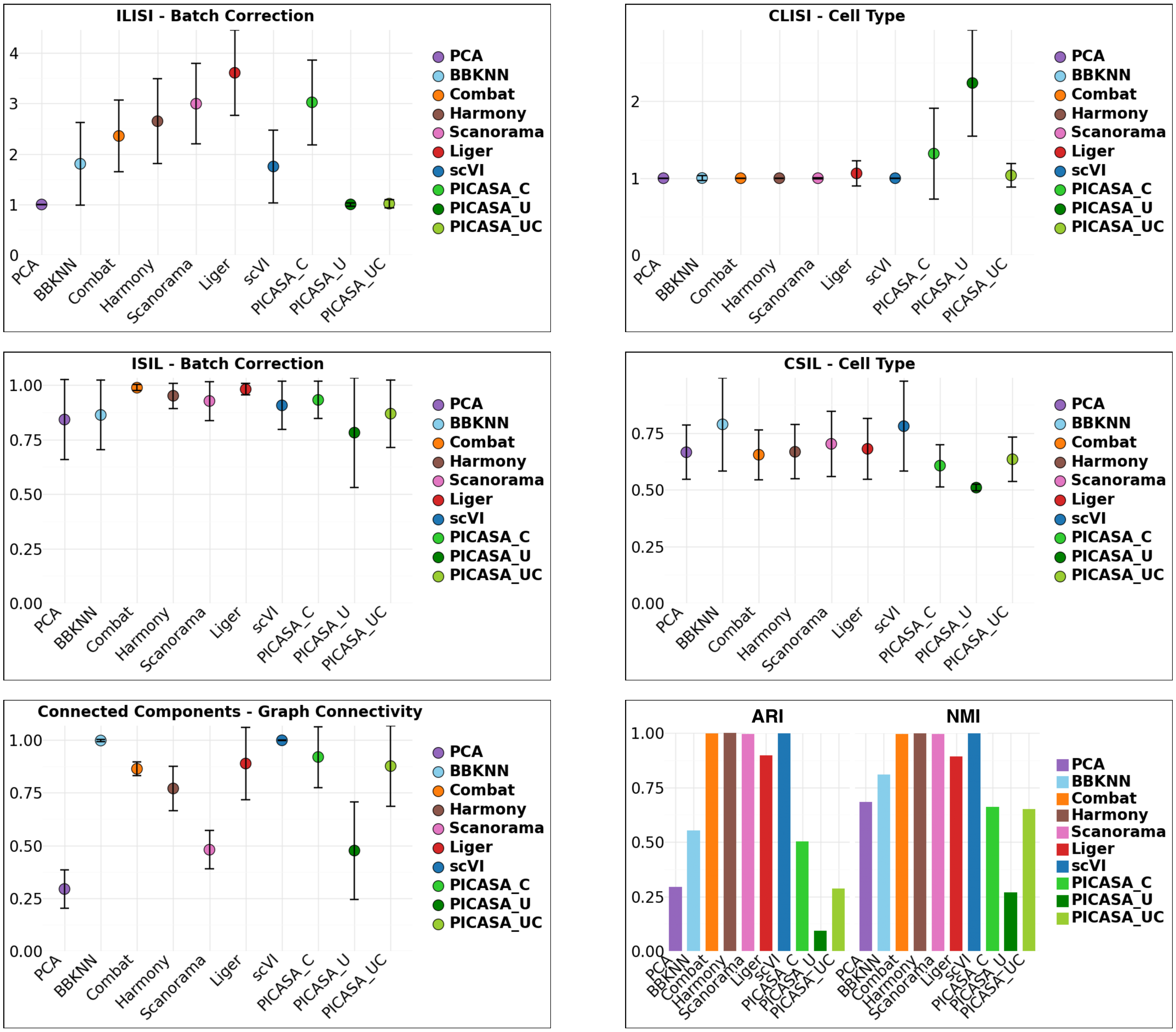

**Fig. S10**: Comparative analysis of PICASA integrating cells across patients in Simulation-3 (large-scale, unbalanced) dataset. The upper panel plot shows batch label Local Inverse Simpson's Index (LISI) (higher value suggests better mixing of cells across samples) and cell type label Local Inverse Simpson's Index (LISI) (lower value suggests better mixing of cells from the same cell type label). The middle panel plot shows batch label scaled Silhouette score (higher value suggests better mixing of cells across samples) and cell type label scaled Silhouette score (higher value suggests better mixing of cells from the same cell type label). The lower panel plot shows cell type label graph connected component analysis (higher value suggests better mixing of cells across samples), and Adjusted Rand Index (ARI) and Normalized Mutual Information (NMI) score (higher value suggests better mixing of cells from the same cell type label).

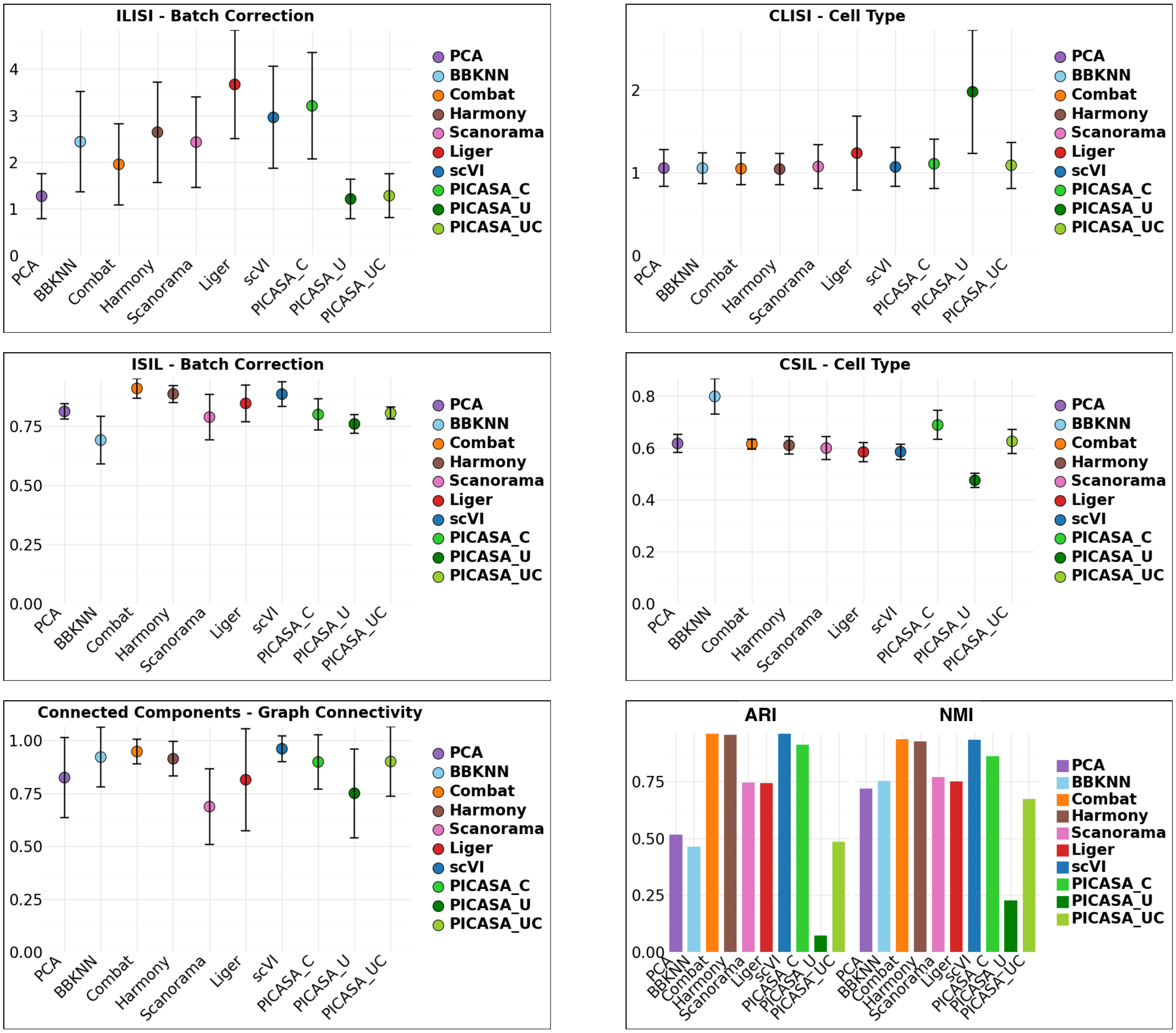

**Fig. S11**: Comparative analysis of PICASA integrating cells across patients in normal pancreas dataset. The upper panel plot shows batch label Local Inverse Simpson's Index (LISI) (higher value suggests better mixing of cells across samples) and cell type label Local Inverse Simpson's Index (LISI) (lower value suggests better mixing of cells from the same cell type label). The middle panel plot shows batch label scaled Silhouette score (higher value suggests better mixing of cells across samples) and cell type label scaled Silhouette score (higher value suggests better mixing of cells from the same cell type label). The lower panel plot shows cell type label graph connected component analysis (higher value suggests better mixing of cells across samples), and Adjusted Rand Index (ARI) and Normalized Mutual Information (NMI) score (higher value suggests better mixing of cells from the same cell type label).

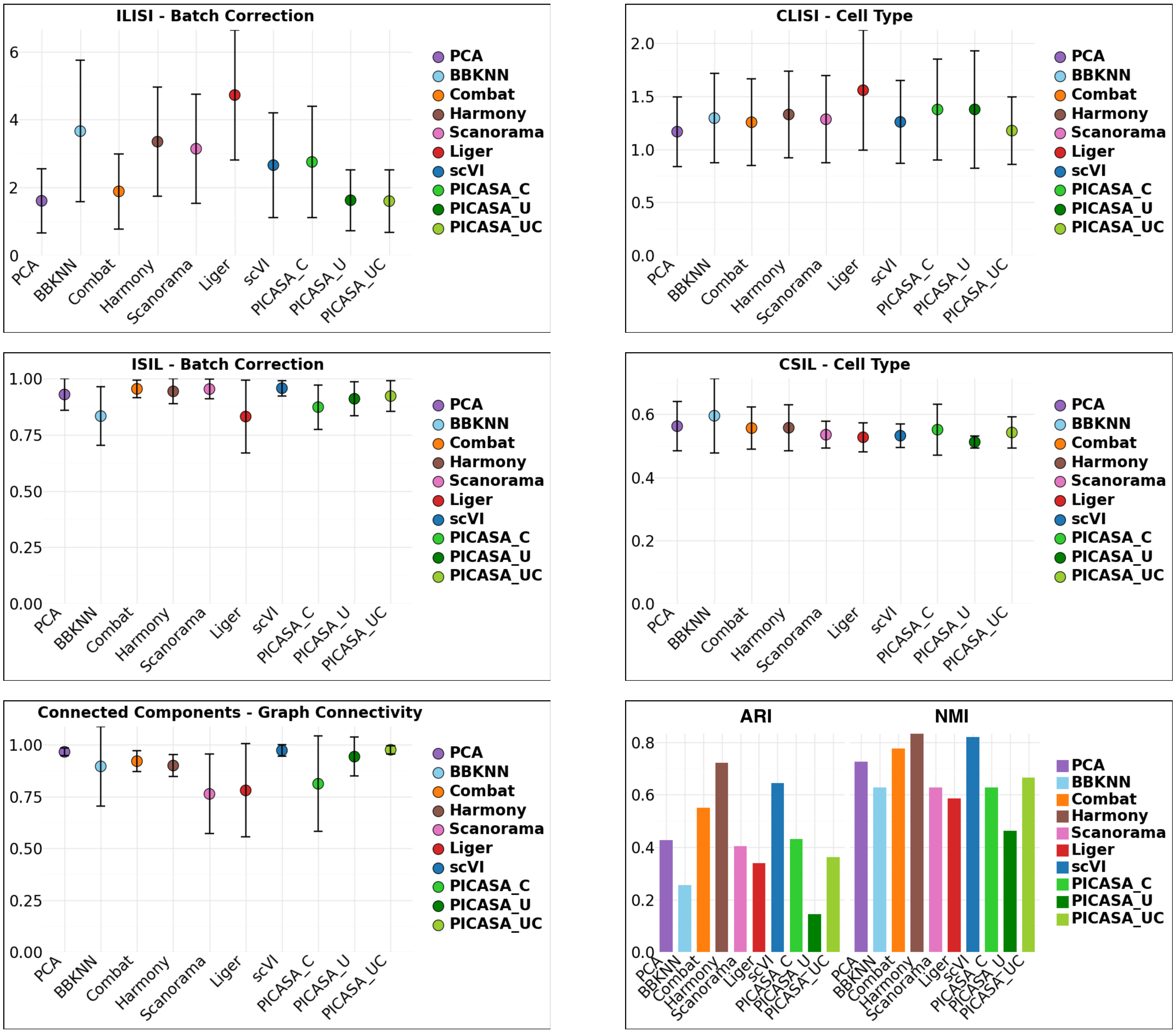

**Fig. S12**: Comparative analysis of PICASA integrating cells across patients in breast cancer dataset. The upper panel plot shows batch label Local Inverse Simpson's Index (LISI) (higher value suggests better mixing of cells across samples) and cell type label Local Inverse Simpson's Index (LISI) (lower value suggests better mixing of cells from the same cell type label). The middle panel plot shows batch label scaled Silhouette score (higher value suggests better mixing of cells across samples) and cell type label scaled Silhouette score (higher value suggests better mixing of cells from the same cell type label). The lower panel plot shows cell type label graph connected component analysis (higher value suggests better mixing of cells across samples), and Adjusted Rand Index (ARI) and Normalized Mutual Information (NMI) score (higher value suggests better mixing of cells from the same cell type label).

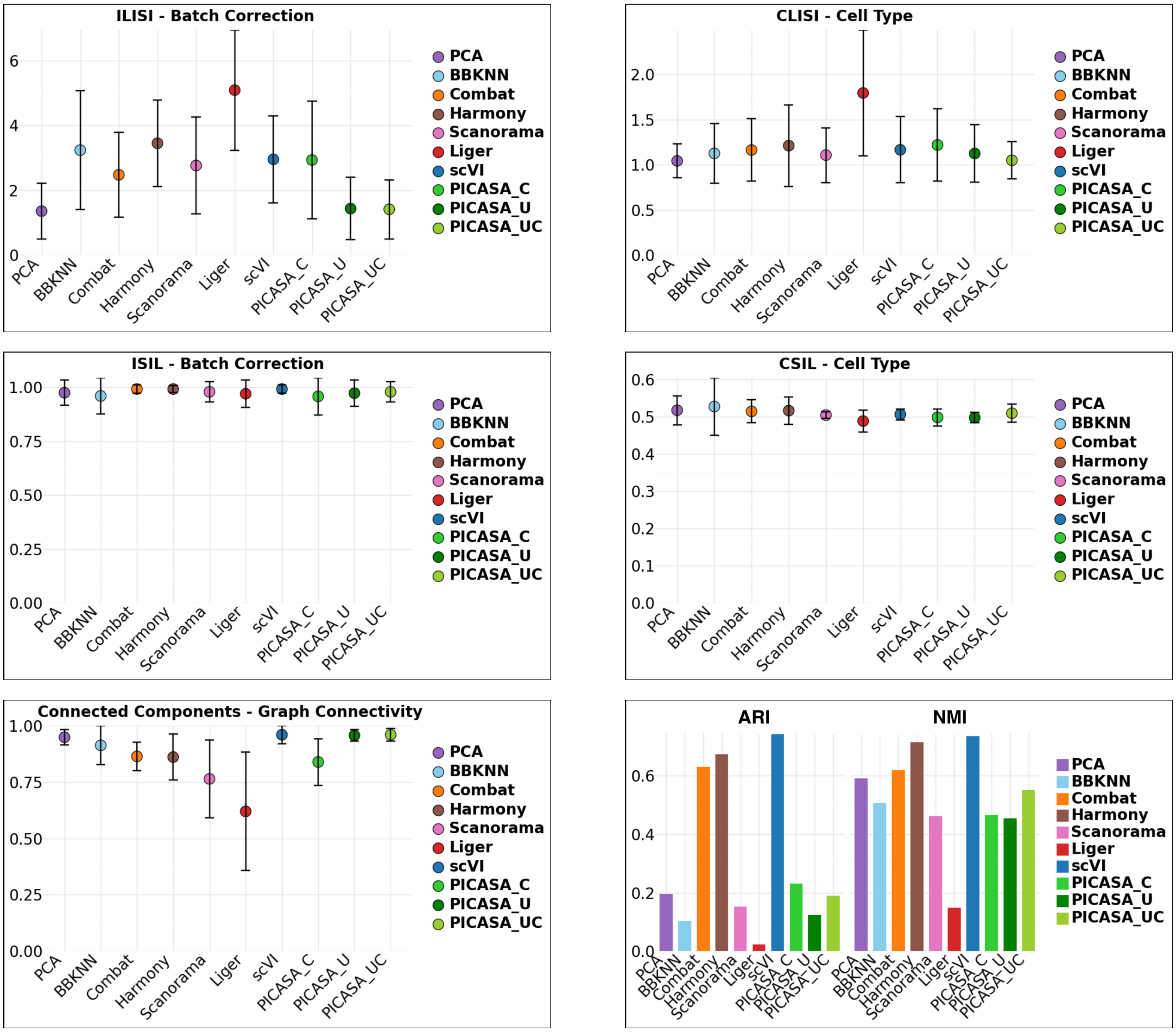

**Fig. S13**: Comparative analysis of PICASA integrating cells across patients in lung cancer dataset. The upper panel plot shows batch label Local Inverse Simpson's Index (LISI) (higher value suggests better mixing of cells across samples) and cell type label Local Inverse Simpson's Index (LISI) (lower value suggests better mixing of cells from the same cell type label). The middle panel plot shows batch label scaled Silhouette score (higher value suggests better mixing of cells across samples) and cell type label scaled Silhouette score (higher value suggests better mixing of cells from the same cell type label). The lower panel plot shows cell type label graph connected component analysis (higher value suggests better mixing of cells across samples), and Adjusted Rand Index (ARI) and Normalized Mutual Information (NMI) score (higher value suggests better mixing of cells from the same cell type label).

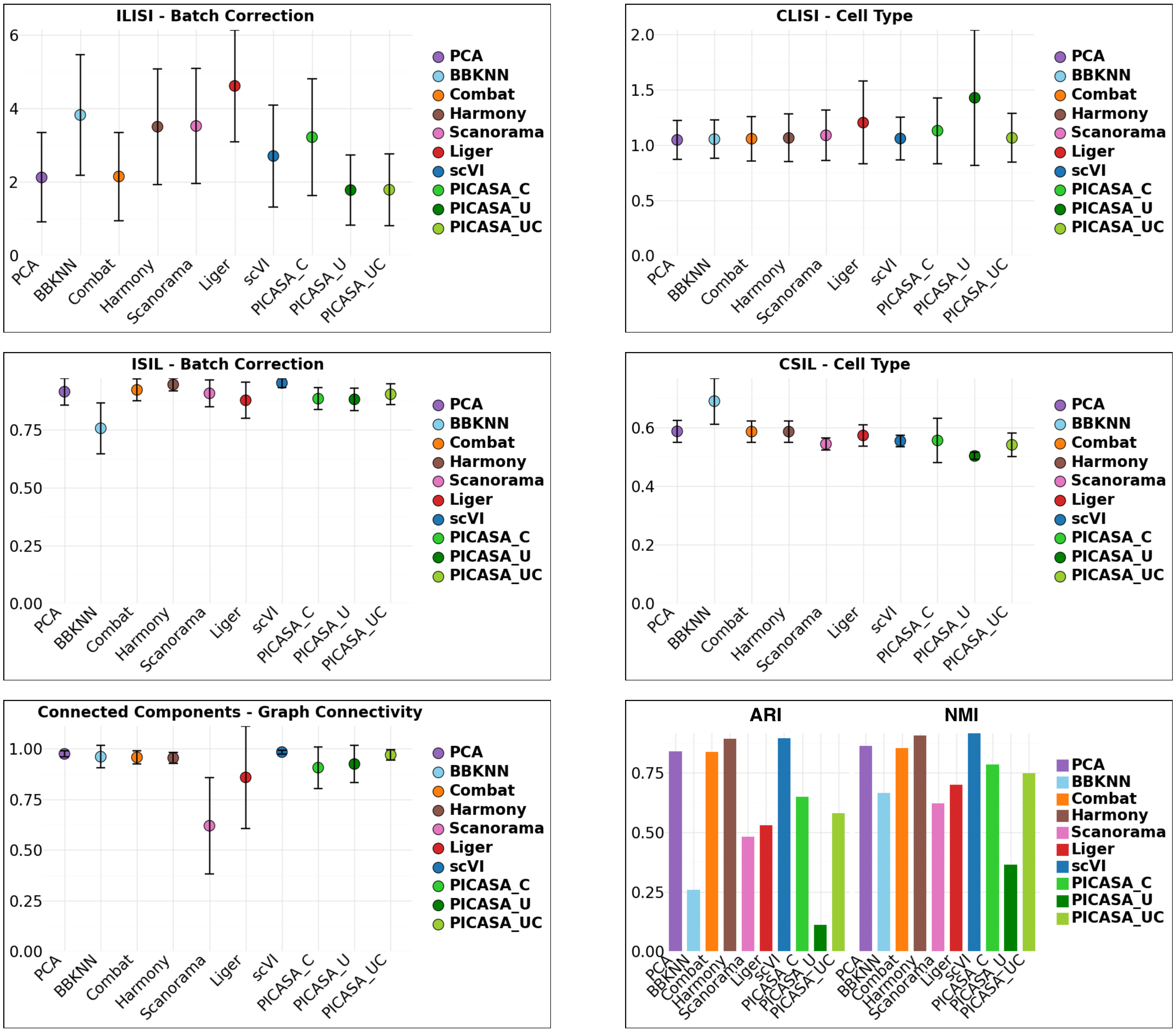

**Fig. S14**: Comparative analysis of PICASA integrating cells across patients in ovarian cancer dataset. The upper panel plot shows batch label Local Inverse Simpson's Index (LISI) (higher value suggests better mixing of cells across samples) and cell type label Local Inverse Simpson's Index (LISI) (lower value suggests better mixing of cells from the same cell type label). The middle panel plot shows batch label scaled Silhouette score (higher value suggests better mixing of cells across samples) and cell type label scaled Silhouette score (higher value suggests better mixing of cells from the same cell type label). The lower panel plot shows cell type label graph connected component analysis (higher value suggests better mixing of cells across samples), and Adjusted Rand Index (ARI) and Normalized Mutual Information (NMI) score (higher value suggests better mixing of cells from the same cell type label).

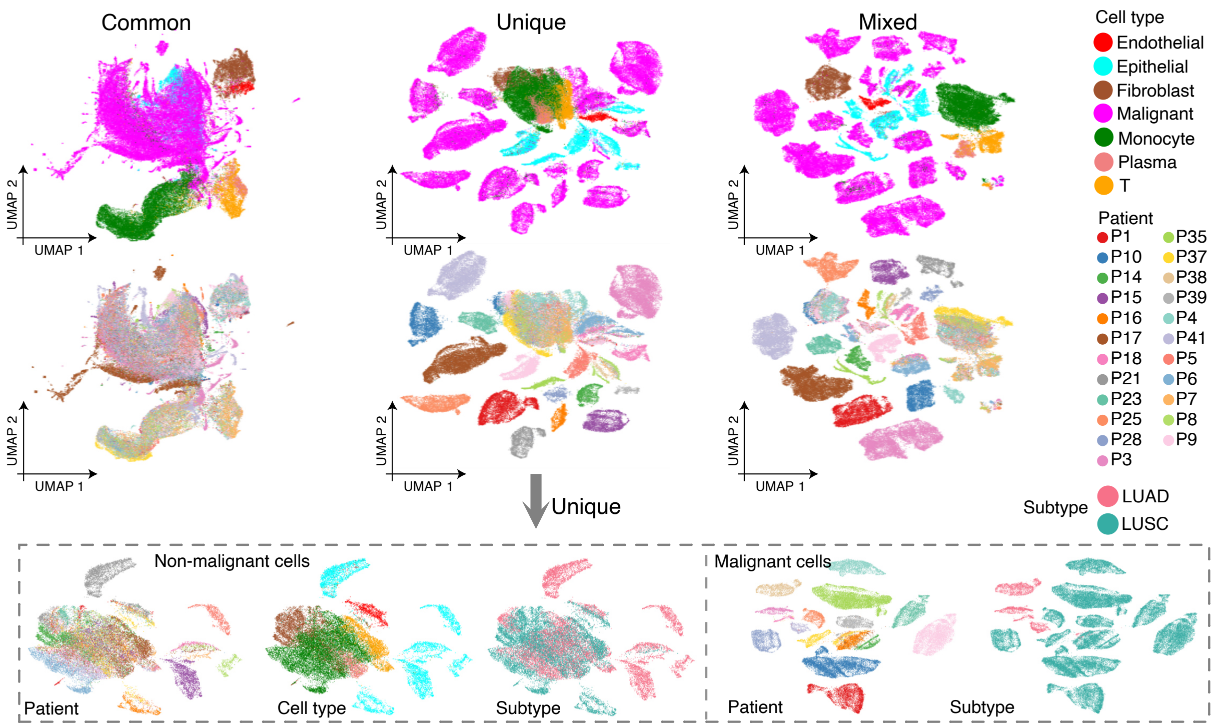

**Fig. S15**: PICASA captures shared cell type and histologically defined cancer subtype-specific effects in lung cancer. UMAP of decomposed common, unique, and mixed representations in lung cancer data. UMAP of cancer and non-cancer cells from unique representation space (lower panel).

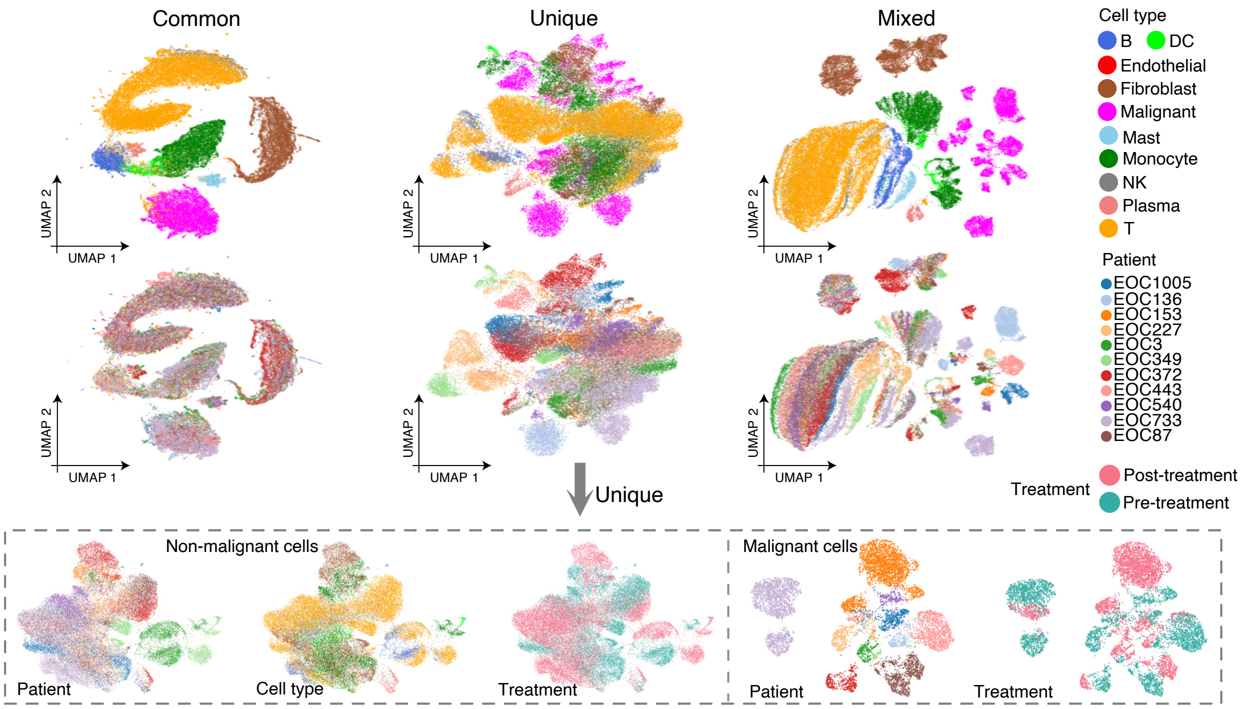

**Fig. S16**: PICASA captures shared cell type and treatment based patient-specific effects in ovarian cancer. UMAP of decomposed common, unique, and mixed representations in ovarian cancer data. UMAP of cancer and non-cancer cells from unique representation space (lower panel).

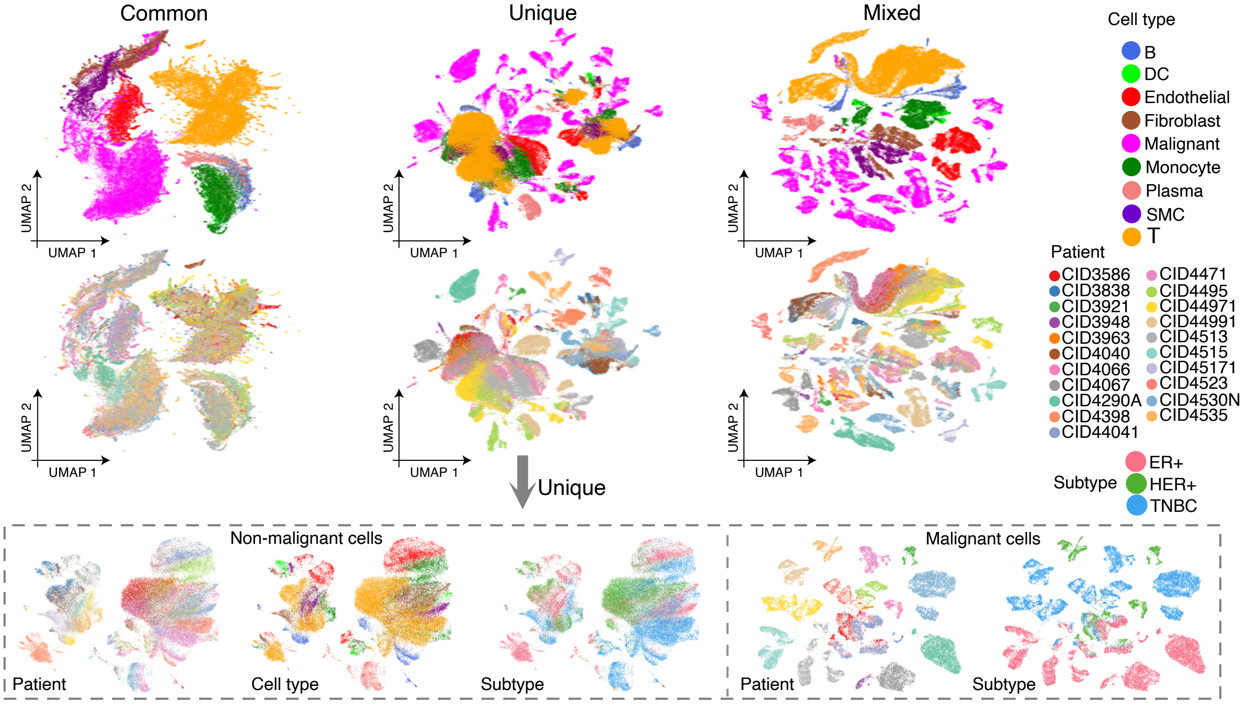

**Fig. S17**: PICASA captures shared cell type and hormone receptor-driven patient-specific effects in breast cancer. UMAP of decomposed common, unique, and mixed representations in breast cancer data. UMAP of cancer and non-cancer cells from unique representation space (lower panel).

### S3 Construction of Matched Cell Pairs

To construct the training dataset for PICSASA, we organized the patient cohort into a sequential pairing schema where each patient is represented in two distinct pairs (e.g., Patient $n$ paired with $n+1$, and Patient $N$ paired back to Patient 1). Once these patient-level pairs were established, we implemented a scalable Approximate Nearest Neighbor (ANN) search using the Annoy library [20] in Python to identify corresponding cell-level pairs. For each target patient in a pair, we constructed a high-dimensional index of the gene expression space using a forest of 50 random projection trees (number_of_trees=50). This index was queried for every cell in the source patient to identify the single most similar counterpart (nbrsize=1) based on an angular distance metric. This metric calculates the cosine similarity between expression vectors, ensuring that the matching is driven by relative biological states rather than technical library size. The resulting pairwise correspondences across the entire cohort allow the model to learn patient-invariant commonalities by identifying shared features between these transcriptionally similar cells.

### S4 Copy Number Variation (CNV) analysis

Copy number variation (CNV) profiles were inferred from single-cell RNA-seq data using the R package CopyKAT [21]. CNV analysis was performed on two types of transcriptomic data: (i) raw single-cell RNA-seq data and (ii) patient-specific expression data estimated from the trained PICASA model after removing shared cell-type effects using a forward generation approach. To minimize potential batch effects, CopyKAT was applied on a patient-by-patient basis within the same cell type for both transcriptomic datasets. Gene symbols were used as identifiers, with a minimum of five genes per chromosome and a sliding window size of 25 genes. CNV calling was performed using a Kolmogorov–Smirnov cutoff of 0.1 and Euclidean distance for hierarchical clustering. Genome-wide CNV profiles were inferred using the hg20 reference genome annotation, and parallel computation was enabled using 32 CPU cores. Identical parameters were used for all analyses to ensure direct comparability between CNV profiles derived from raw and patient-specific transcriptomic data.

### S5 Computational architecture and scalability

The PICASA model adopts a simplified transformer-based architecture in which cross-attention is applied to embeddings derived from the 2,000 most highly variable genes, enabling efficient modelling of gene–gene interactions while controlling computational complexity (Tables S1, S2, S3). The commonality network operates on gene embeddings of dimension 3000 and applies a single attention module with query, key, and value dimensions of size 25. The encoder in the commonality network is a two-layer multilayer perceptron with hidden layer sizes of 100 and 25, producing a common latent representation of dimension 25, followed by a projection network composed of two layers of size 50 each. The patient-specific (unique) network takes the entire raw transcriptomic data ~25K genes and the common latent representation trained from the commonality network as input and uses an encoder with two fully connected layers of sizes 128 and 25 to learn a patient-specific latent space of dimension 25, followed by a decoder with two fully connected layers of size 128 each to reconstruct patient-specific expression patterns. Training is performed with batch sizes of 100 and 128 for the commonality and unique networks, respectively, and learning rates of $1\times10$   and $1\times10^{3}$. Training was conducted on a single GPU with 25 GB of memory and processed approximately at a rate of 100,000 cells per hour. Rather than aiming to train large-scale foundation models, we designed a lightweight architecture consisting of a single transformer layer with one attention head and a reduced feature set of highly variable genes, demonstrating that such a simplified model is sufficient to capture general cell-type signatures while substantially lowering computational cost. With the availability of computational resources, model capacity can be increased by incorporating additional transformer layers, attention heads, and a larger number of gene features to approach foundation-model–level performance. Scalability can be further enhanced through gene symbol–based embedding and tokenization strategies, as well as by adopting more memory-efficient attention implementations, thereby reducing both memory footprint and computational overhead.

Table S1: Hyperparameters for PICASA model.

| Hyperparameter | Default | Description |
|---|---|---|
| pair__mode | seq | The mode to pair datasets is [seq] for sequential pairing datasets such that before and after pairs share a dataset, [dist] for calculating truncated SVD for each dataset, measuring Euclidean distances among datasets, and pairing the closest datasets, and [all] for pairing all datasets with each other. |
| estimate__neighbour | approx | The method to estimate cell pairs from two datasets in a pair is [approx] for using ANNOY method and [exact] for exact Euclidean distance using the cdist function in scipy. |
| common__net: batch__size | 64 | The batch size for training commonality network. |
| common__net: embedding__dim | 1000 | The embedding size for gene features. |
| common__net: attention__dim | 10 | The size of the attention module dimension. This is the same for query, key, and value matrices. |
| common__net: encoder__layers | [100,10] | The architecture of encoder in commonality network. |
| common net: latent__dim | 10 | The size of latent dimension for commonality latent space. |
| common__net: projection__layers | [25,25] | The architecture of the projection layer in a commonality network. |
| common__net: learning rate | 1.00E-05 | The learning rate for commonality network. |
| common__net: pair__importance__weight | 0 | The value between 0 and 1 guides the commonality network to give more importance to matched gene pairs during training attention parameters. |
| common__net:corruption__tol | 10 | The standard deviation value to mask corrupt cell pairs with very high distance values. |
| common__net: cl__loss__mode | none | The method to calculate contrastive loss is [none] for InfoNCE base loss (see methods), [rare] for adding importance to rare cell types, [wclust] for increasing distance among negative pairs, and [margin] for adding importance to separate hard examples of negative pairs. |
| common__net: epochs | 1 | The number to iterate a pair of dataset. |
| common__net: meta__epochs | 10 | The number to iterate through all pairs of datasets. |
| common__net: eval__batch__size | 500 | The size of the batch to use for the inference step to estimate commonality latent space for all datasets. |
| unique__net: encoder__layers | [128,10] | The architecture of the encoder in a patient-specific network. |
| unique__net: common__latent__dim | 10 | The dimension size of commonality latent space from a commonality network. |
| unique__net: latent__dim | 10 | The size of the latent dimension for a patient-specific latent space. |
| unique__net: decoder__layers | [128,128] | The size of the decoder layer in a patient-specific network. |
| unique__net: learning__rate | 1.00E-03 | The learning rate for a patient-specific network. |
| unique__net: batch__size | 128 | The batch size for training a patient-specific network. |
| unique__net: epochs | 250 | The number to iterate for training patient-specific network. |
| unique__net:eval__batch__size | 1000 | The number of data size for inference in a patient-specific network. |

Table S2: Hyperparameters for normal datasets used in PICASA model.

| Hyperparameter | Sim1 | Sim2 | Sim3 | Pancreas |
|---|---|---|---|---|
| pair__mode | seq | seq | seq | seq |
| estimate__neighbour | approx | approx | approx | approx |
| common__net: batch__size | 100 | 100 | 100 | 100 |
| common__net: embedding__dim | 1000 | 2000 | 2750 | 3000 |
| common__net: attention__dim | 15 | 15 | 15 | 15 |
| common__net: encoder__layers | [100,15] | [100,15] | [100,15] | [100,15] |
| common net: latent__dim | 15 | 15 | 15 | 15 |
| common__net: projection__layers | [25,25] | [25,25] | [25,25] | [25,25] |
| common__net: learning rate | 1.00E-03 | 1.00E-05 | 1.00E-03 | 1.00E-03 |
| common__net: pair__importance__weight | 0.1 | 0.5 | 0.75 | 1 |
| common__net:corruption__tol | 10 | 10 | 10 | 10 |
| common__net: cl__loss__mode | none | none | none | none |
| common__net: epochs | 1 | 1 | 1 | 1 |
| common__net: meta__epochs | 5 | 15 | 15 | 15 |
| common__net: eval__batch__size | 500 | 500 | 500 | 500 |
| unique__net: encoder__layers | [128,15] | [128,15] | [128,15] | [128,15] |
| unique__net: common__latent__dim | 15 | 15 | 15 | 15 |
| unique__net: latent__dim | 15 | 15 | 15 | 15 |
| unique__net: decoder__layers | [128,128] | [128,128] | [128,128] | [128,128] |
| unique__net: learning__rate | 1.00E-03 | 1.00E-03 | 1.00E-03 | 1.00E-03 |
| unique__net: batch__size | 128 | 128 | 128 | 128 |
| unique__net: epochs | 250 | 250 | 250 | 250 |
| unique__net:eval__batch__size | 1000 | 1000 | 1000 | 1000 |

Table S3: Hyperparameters for cancer datasets used in PICASA model.

| Hyperparameter | Lung Cancer | Ovarian Cancer | Breast Cancer |
|---|---|---|---|
| pair_mode | seq | seq | seq |
| estimate_neighbour | approx | approx | approx |
| common_net: batch_size | 100 | 100 | 100 |
| common_net: embedding_dim | 3000 | 3000 | 3000 |
| common_net: attention_dim | 25 | 25 | 25 |
| common_net: encoder_layers | [100,25] | [100,25] | [100,25] |
| common net: latent_dim | 25 | 25 | 25 |
| common_net: projection_layers | [50,50] | [50,50] | [50,50] |
| common_net: learning rate | 1.00E-05 | 1.00E-05 | 1.00E-05 |
| common_net: pair_importance_weight | 0.75 | 0.75 | 0.75 |
| common_net:corruption_tol | 10 | 10 | 10 |
| common_net: cl_loss_mode | none | none | none |
| common_net: epochs | 1 | 1 | 1 |
| common_net: meta_epochs | 15 | 15 | 15 |
| common_net: eval_batch_size | 500 | 500 | 500 |
| unique_net: encoder_layers | [128,25] | [128,25] | [128,25] |
| unique_net: common_latent_dim | 25 | 25 | 25 |
| unique_net: latent_dim | 25 | 25 | 25 |
| unique_net: decoder_layers | [128,128] | [128,128] | [128,128] |
| unique_net: learning_rate | 1.00E-03 | 1.00E-03 | 1.00E-03 |
| unique_net: batch_size | 128 | 128 | 128 |
| unique_net: epochs | 250 | 250 | 250 |
| unique_net:eval_batch_size | 1000 | 1000 | 1000 |

### S6 Cross-attention module reveals gene interactions in normal and cancer datasets

We further investigated the biological interpretability of gene interaction patterns learned by the cross-attention module using the normal pancreas and cancer datasets. The attention matrix learned by the model shows distinct patterns of gene interactions for each cell type (Figure S18) present in the datasets. These interaction scores can be further investigated to extract biological characteristics of the shared cell types. We confirmed that the gene interaction signals learned by the model are indeed biologically pertinent by querying interaction patterns of known cell type markers established by the previous studies.

#### S6.1 Normal pancreas single-cell data:

For pancreas dataset, the marker genes for each cell type were among the highly active genes in the interaction matrix of the corresponding clusters (Figure S19). For example, active markers for integrated cell types for pancreas included - alpha (*PLCE1*, *LOXL4*), beta (*NPTX2*, *DLK1*), delta (*LEPR*, *RBP4*), acinar (*PLA2G1B*, *CPA1*), and ductal (*KRT19*, *SPP1*).

To solidify our observation that integrated cell clusters represent biological cell types shared among batches, we investigated the gene interaction patterns of all highly variable genes within each cell type. We ranked interaction scores and performed gene set enrichment analysis against the PanglaoDB database (among the top 5 pathways with gene set size of min 10 and max 500) [22]. The ranked gene list estimated by the model for each cell type was significantly associated with curated gene markers belonging to similar cell types and their origin (Figure S20). For example, all the major cell types in the pancreas - acinar ($p<0.001$), alpha ($p=0.03$), beta ($p=0.001$), delta ($p=0.027$), gamma ($p=0.212$), ductal ($p<0.001$), and endothelial cells ($p<0.001$) were associated with matched cell type in the PanglaoDB database.

#### S6.2 Breast cancer:

We repeated the gene interaction analysis using the attention matrix in breast cancer dataset as well. The cancer dataset [23] consists of 21 patients clinically stratified in three cancer subtypes - 8 Luminal (*ER*+, *PR*+/−), 5 *HER2*+ (*HER2*+, *ER*+/−, *PR*+/−) and 8 triple negative (TNBC; *ER*−, *PR*−, *HER2*−). PICASA isolated shared cell type-specific clusters in the common space, where integrated clusters represented all the major cell types in the breast tumour microenvironment (Figure S17). First, we generated cell clusters based on the shared space representing all cell types across patients in the TME: clusters C8 for endothelial, C1 for monocyte, C18 for plasma, C4 for T, C11 for B, C15 for DC, C0 for malignant, and C10 for fibroblast cell types (Figure S22). We identified known marker genes among the genes with high scores in the model-estimated cross-attention interaction matrix (Figure S23). For example, active markers for integrated clusters of breast cancer cells included - endothelial (*PECAM1*, *CD34*), fibroblasts (*PDGFRB*, *COL1A1*), malignant (*EPCAM*, *MKI67*), monocyte (*CD68*, *CD14*), plasma (*JCHAIN*, *IGKC*), and lymphoid cells: B (*CD27*, *IGHD*) and T (*CD3D*, *TIGIT*) (Figure S21).

For cancer cells, *EPCAM* marker gene is highly expressed and a potential target for gene therapy [24]. We found that *IGKC* is also active in cancer cells and is known to be expressed in tumour-infiltrating plasma cells [25]. For cancer-associated fibroblasts, two marker genes, *PDGFRB* and

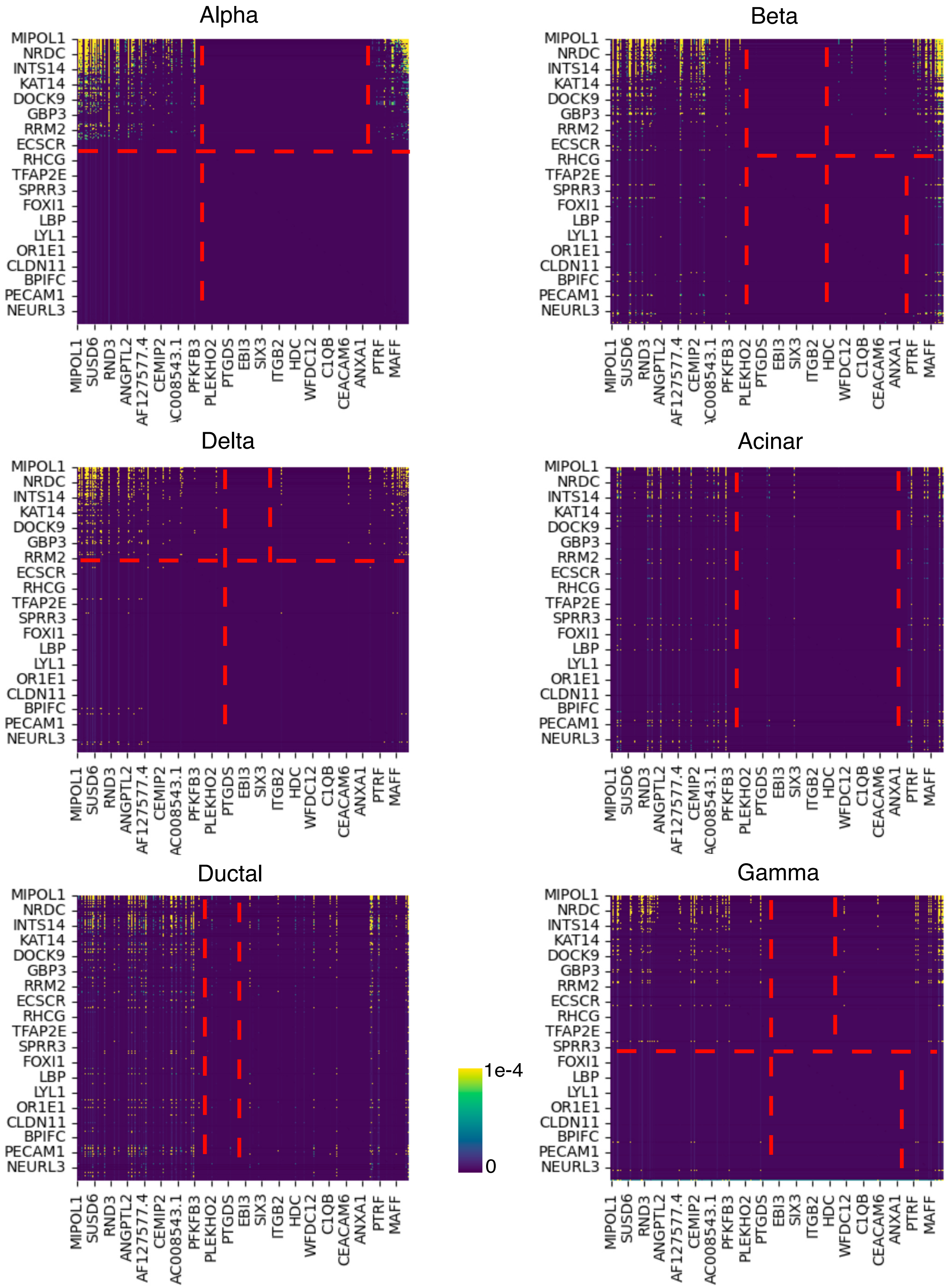

**Fig. S18**: Cross-attention across all 2000 highly variable genes from matched cell pairs shows cell type-specific gene interaction patterns (normal pancreas datasets).

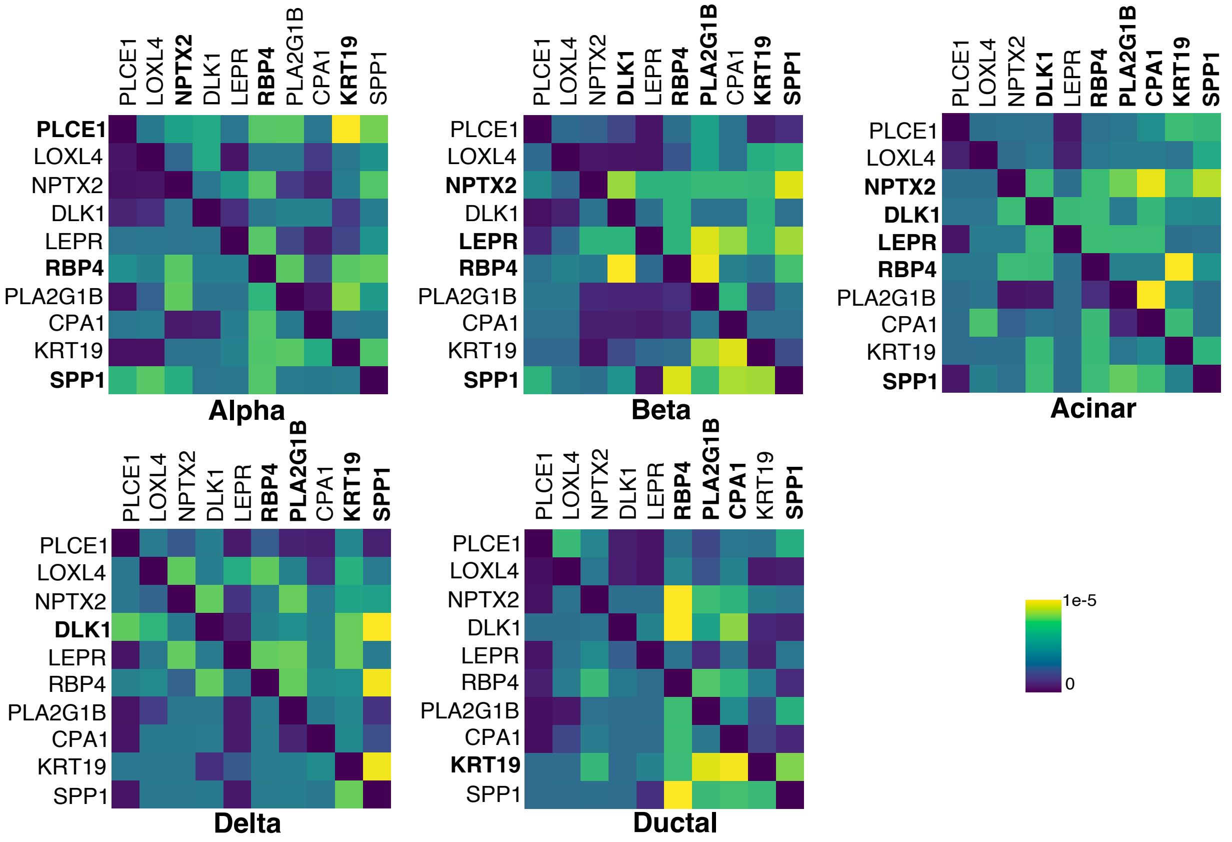

**Fig. S19**: Heatmap of gene interaction scores of known marker genes for different cell types in the normal pancreas data estimated by the cross-attention module.

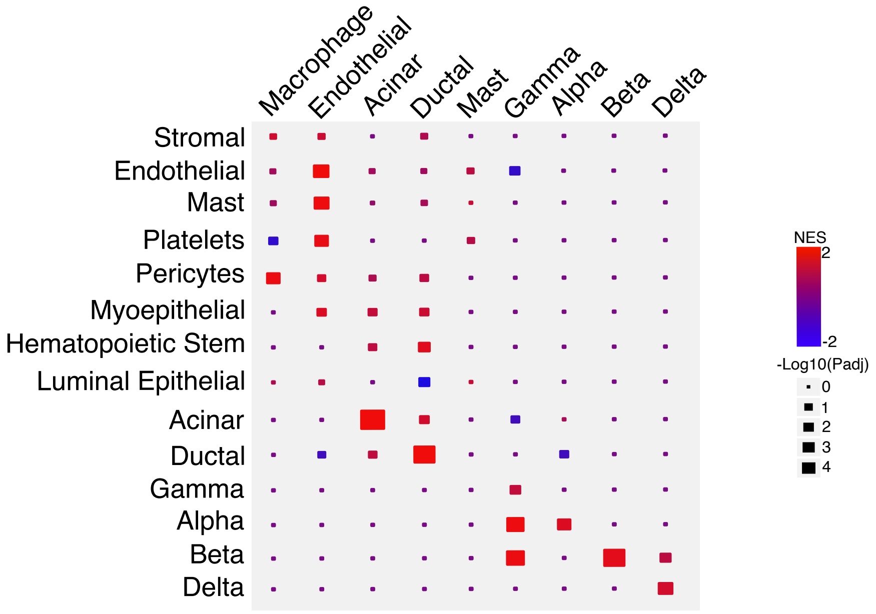

**Fig. S20**: Gene set enrichment analysis for the normal pancreas data based on ranked genes in the cross-attention matrix against cell type markers in the PanglaoDB database.

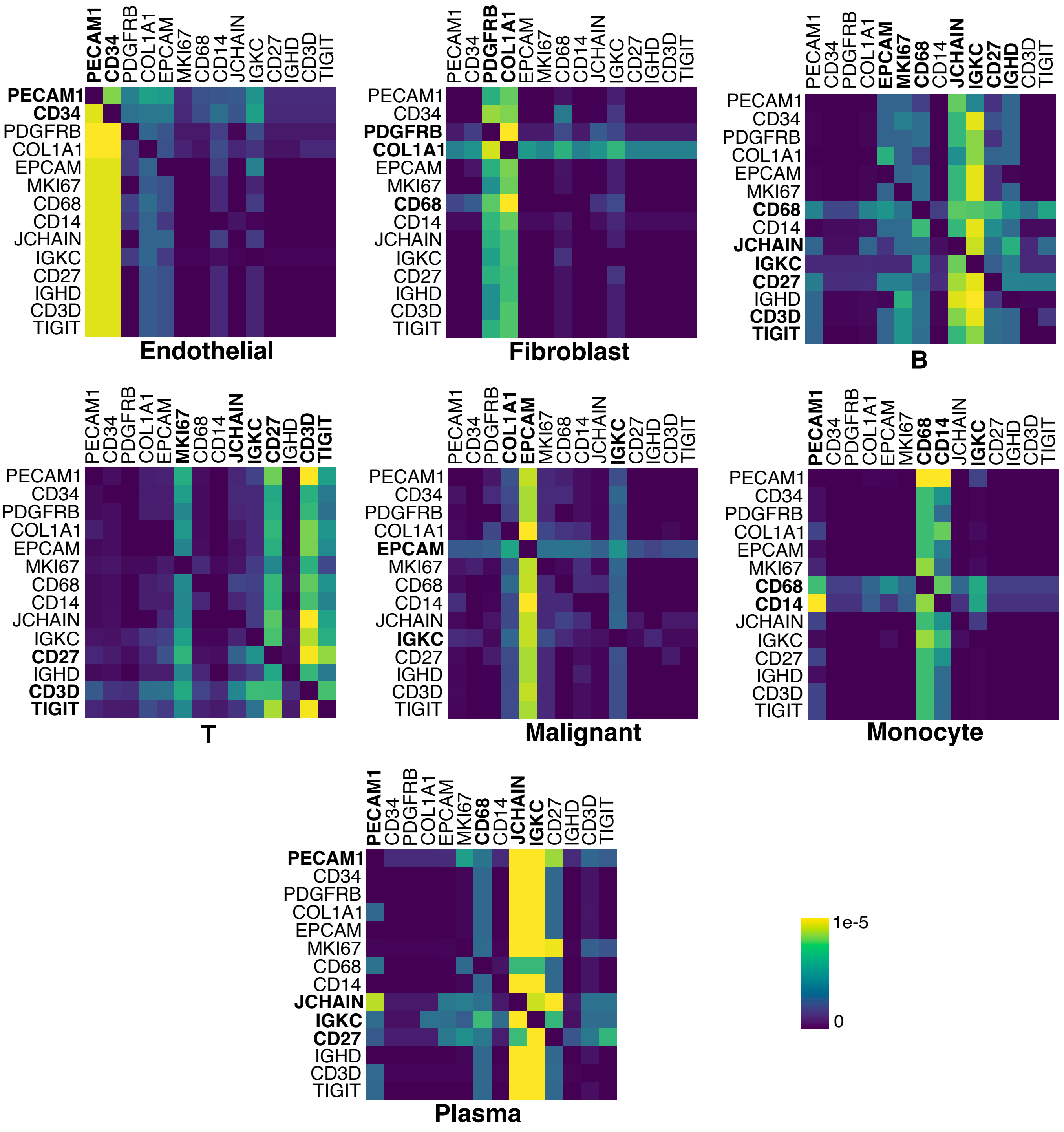

**Fig. S21**: Heatmap of gene interaction scores of known marker genes for different cell types in the breast cancer dataset learned by the cross-attention module.

*COL1A1*, tend to be co-expressed in our model, which is known to form a *COL1A1-PDGFRB* fusion transcript in cancer cells [26]. For the immune cell types, the cross-attention mechanisms show known marker genes interacting with other marker genes across cell types. For monocytes, we found a strong connection between *CD14* and *CD68*. The high activity of these genes suggests a higher level of M2-like macrophages and an aggressive tumour microenvironment [27]. Moreover, the interactions of *JCHAIN* and *IGKC* marker genes in B cells, along with other immune markers, suggest humoral immune responses in the tumour microenvironment [28].

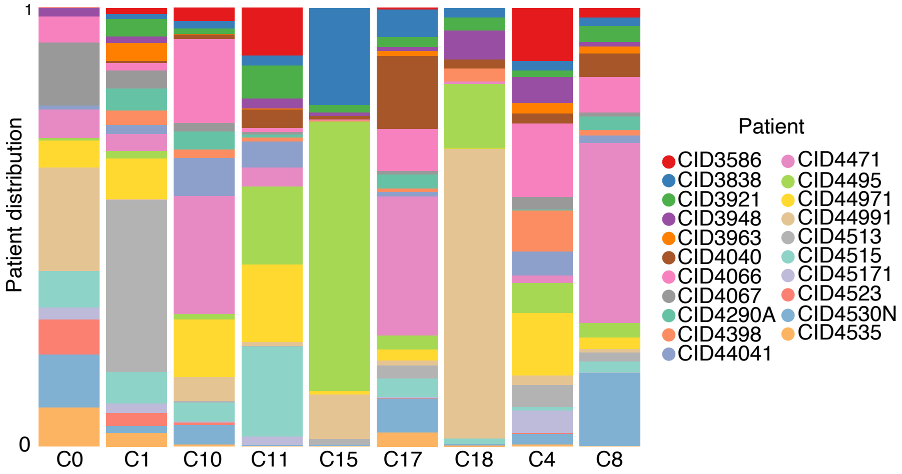

**Fig. S22**: Clusters (Leiden algorithm) in the common space representing shared cell types show the integration of cells across patients in breast cancer data.

The gene set enrichment analysis based on ranking all 2000 highly variable genes in each common cluster was significantly associated with similar cell types in the PanglaoDB database (Figure S23). Here, C0/malignant was associated with luminal epithelial cells ($p<0.001$) and mammary epithelial cells ($p=0.009$). C1/monocyte was associated with monocytes ($p<0.001$), macrophages ($p<0.001$), and microglia ($p<0.001$). C18/plasma was associated with plasma cells ($p=0.005$) and B cells ($p=0.03$). C4/T was associated with T cells ($p=0.014$), natural killer T cells ($p=0.03$), and T regulatory cells ($p=0.05$). C11/B was associated with B cells ($p=0.027$) and plasma Cells ($p=0.003$). C15/DC was associated with monocytes ($p=0.003$) and dendritic cells ($p=0.03$). C8/endothelial was associated with endothelial Cells($p<0.001$). C10/fibroblasts was associated with fibroblasts ($p<0.001$) and myofibroblasts ($p<0.001$).

#### S6.3 Ovarian cancer:

For epithelial ovarian cancer (EOC) cells, two marker genes, *EPCAM* and *WFDC2* tend to be strongly co-expressed/interacting with each other. For HGSOC, EpCAM is a well-known marker gene, particularly highly expressed in chemotherapy-resistant tumour samples [29]. *WFDC2* (or *HE4*) is also another well-established marker gene of ovarian cancer [30], exhibiting albeit subtype specificity [31]. For cancer-associated fibroblasts (CAF), two marker genes, *PDPN* and *DCN*, tend to be co-expressed in our model, strongly aligned with CAF studies in pancreatic ductal adenocarcinoma [32]. For the immune cell types, we found that the model parameters corresponding to marker genes are highly activated–*CD14* for monocytes, *CD8A* and *PTPRC*/CD45 for T-cells, and *MS4A1*/CD20 along with *CD79A* (B-cell receptor complex) for B-cells. The cross-attention mechanisms link these marker genes with other marker genes. For monocytes, we found a strong

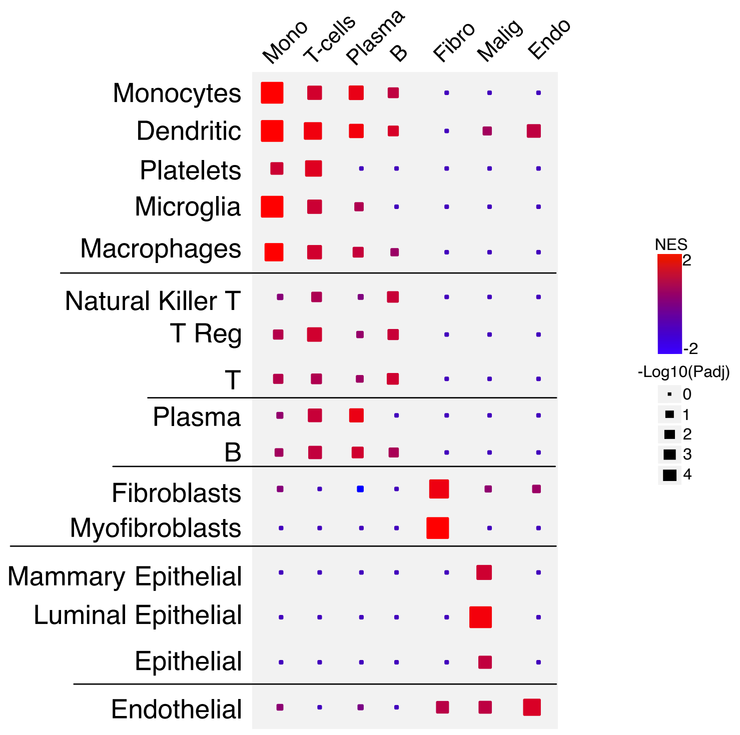

**Fig. S23**: Gene set enrichment analysis of breast cancer data based on ranked genes in the cross-attention matrix with the known cell-type marker genes in the PanglaoDB database.

connection between *CD14* and *THBD* (Fig. 3C). As *THBD* is specifically expressed in hypoxic tumour regions [33], the high activity of *CD14* together with *THBD* suggests a dominant role M2 macrophages for this HGSOC cohort. Moreover, the co-occurrence of *FCER1G* corroborates that the monocytes/macrophages are highly tumour-infiltrating [34], showing clear pathological characteristics of tumour microenvironments.

#### S6.4 Lung cancer:

The immune cell types enrich well-known cell surface and other marker genes/proteins: a surface marker gene *CD14* and lysosome (*LYZ*) gene for monocytes, immunoglobulin genes (*IGHG3* and *JCHAIN*) for plasma cells, and *CD2* and *IGHG3* for T-cells. Moreover, for endothelial cells, a key autocrine regulator gene, *ANGPT2* (angiopoietin-2), is highly expressed [35] and interacts with a wide spectrum of genes, including extracellular matrix protein genes, *COL1A2* and *DCN2*, which are more pertinent to epithelial and fibroblasts. *NAPSA* (napsin A) is highly coexpressed with *KRT8A* and *LYZ* in the epithelial cells' patterns, as implicated in an independent experimentally validated study [36]. We notice a switching pattern of *NAPSA* and *KRT6A* expressions between the epithelial and malignant cells. Considering that *KRT6A* promotes epithelial-to-mesenchymal transition (EMT) in many cancer types [37], including NSCLC, we can speculate systematic rearrangement of gene-gene interaction networks during cancer progression. On the other hand, *DCN* (decorin), an extracellular matrix protein/gene, suppresses the EMT process and functions as a tumour suppressor [38,39]. We found *DCN* is specifically interacting with the other extracellular matrix protein/gene, *COL1A2*, and coexpressed with *KRT6A*, perhaps inhibiting *KRT6A*'s function in EMT, maintaining the fibroblasts non-cancerous.

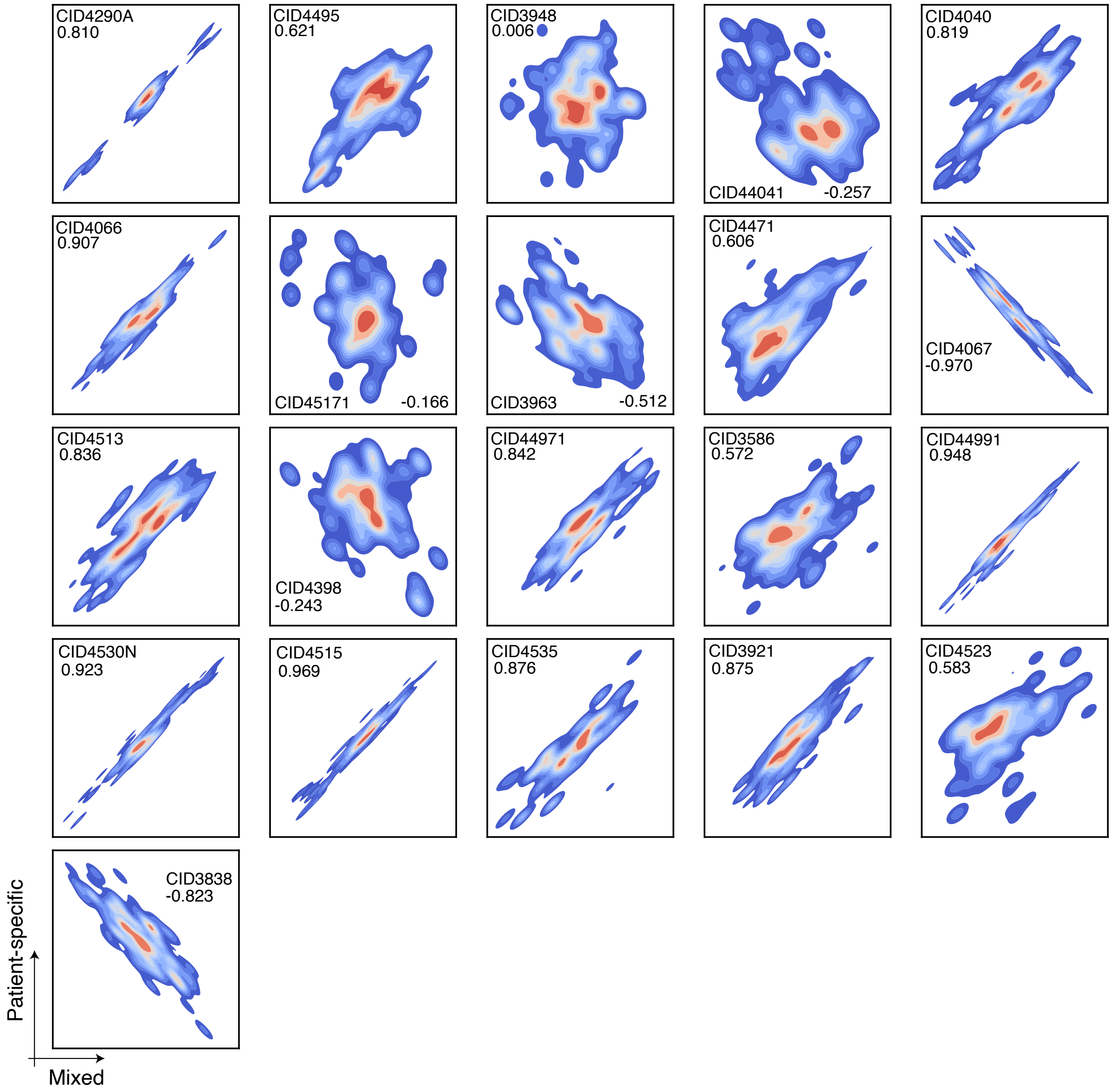

**Fig. S24**: Patient-wise contour plots of CNV profiles using `copyKAT` from patient-specific and mixed representation of breast cancer data (value is Spearman's correlation).

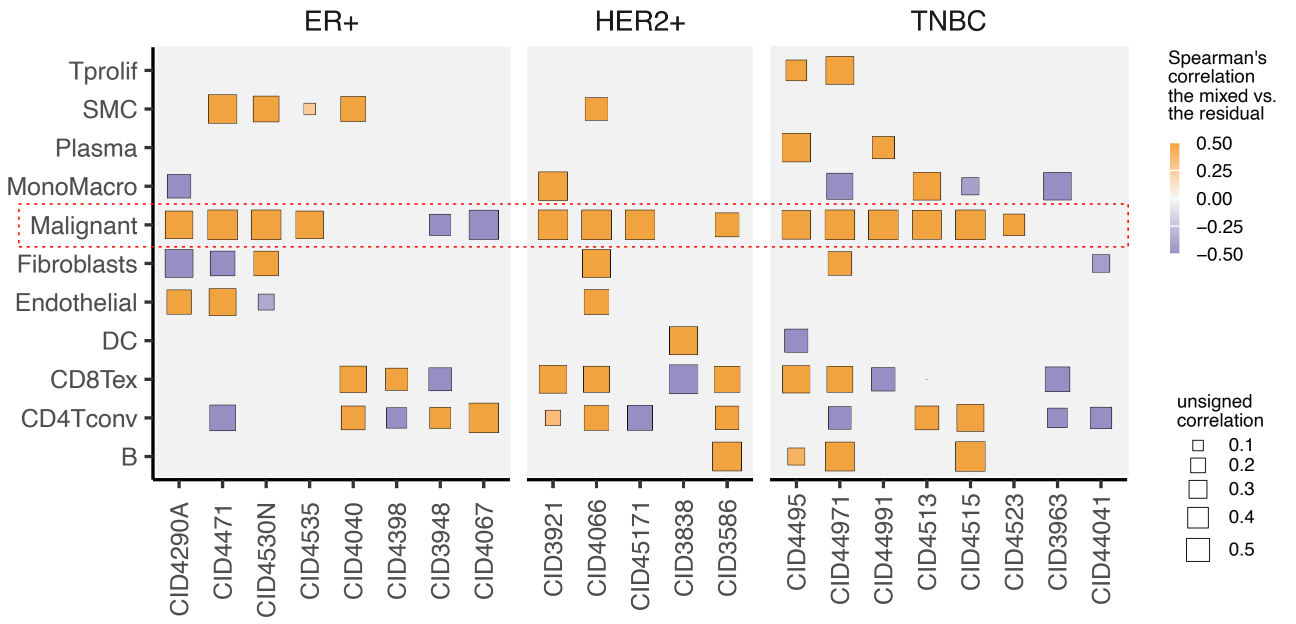

**Fig. S25**: Heatmap of correlation between CNV profiles from patient-specific and mixed representations for different cell types for breast cancer data.

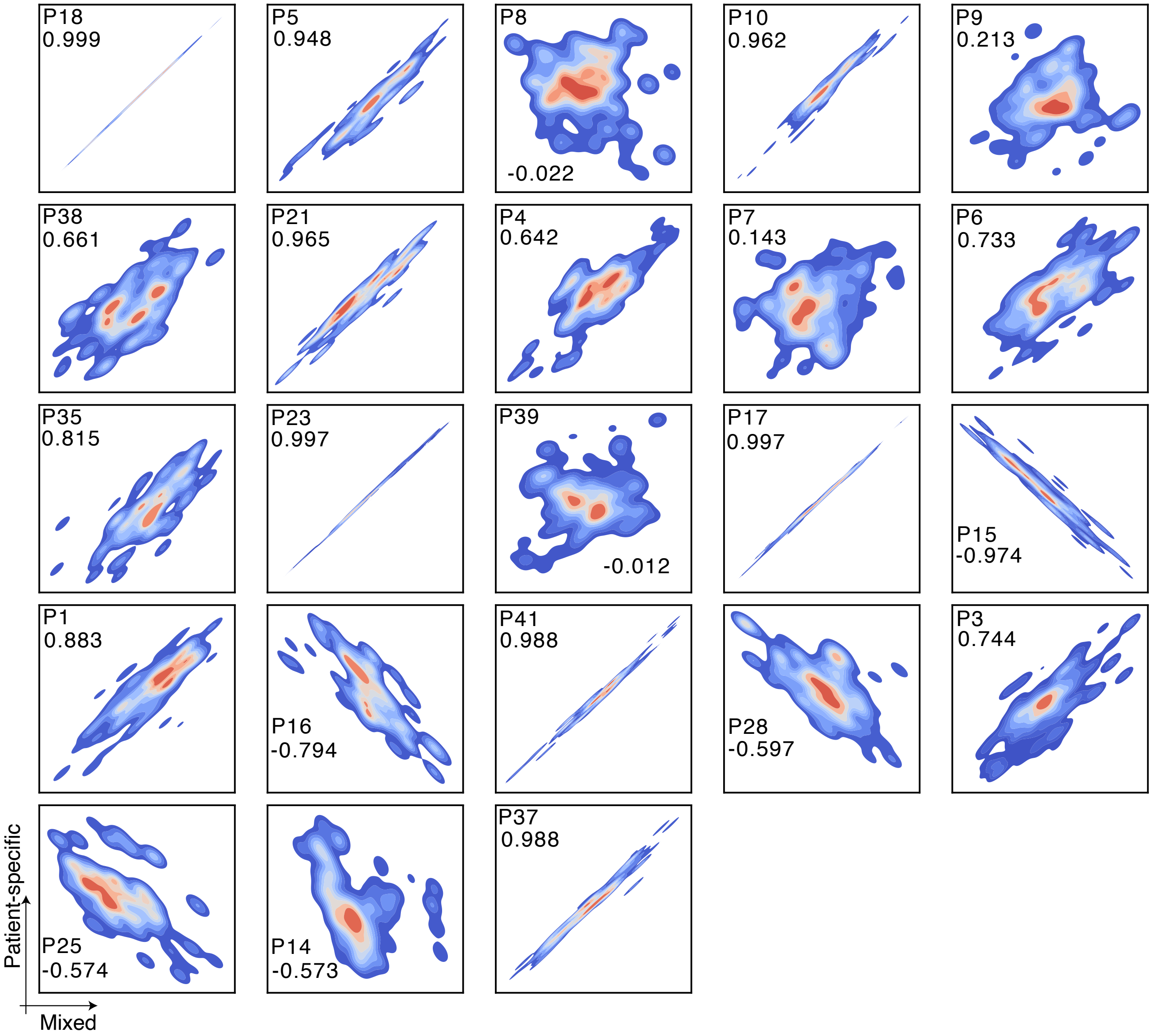

**Fig. S26**: Patient-wise contour plots of CNV profiles using `copyKAT` from patient-specific and mixed representations of lung cancer data (value is Spearman's correlation).

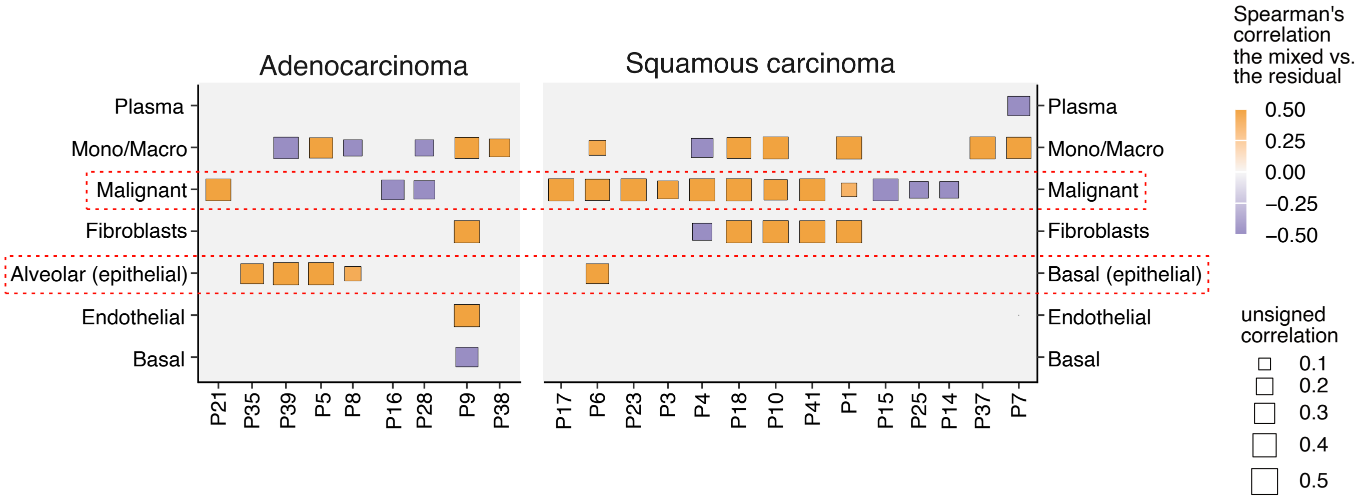

**Fig. S27**: Heatmap of correlation between CNV profiles from patient-specific and mixed representations for different cell types for lung cancer data.

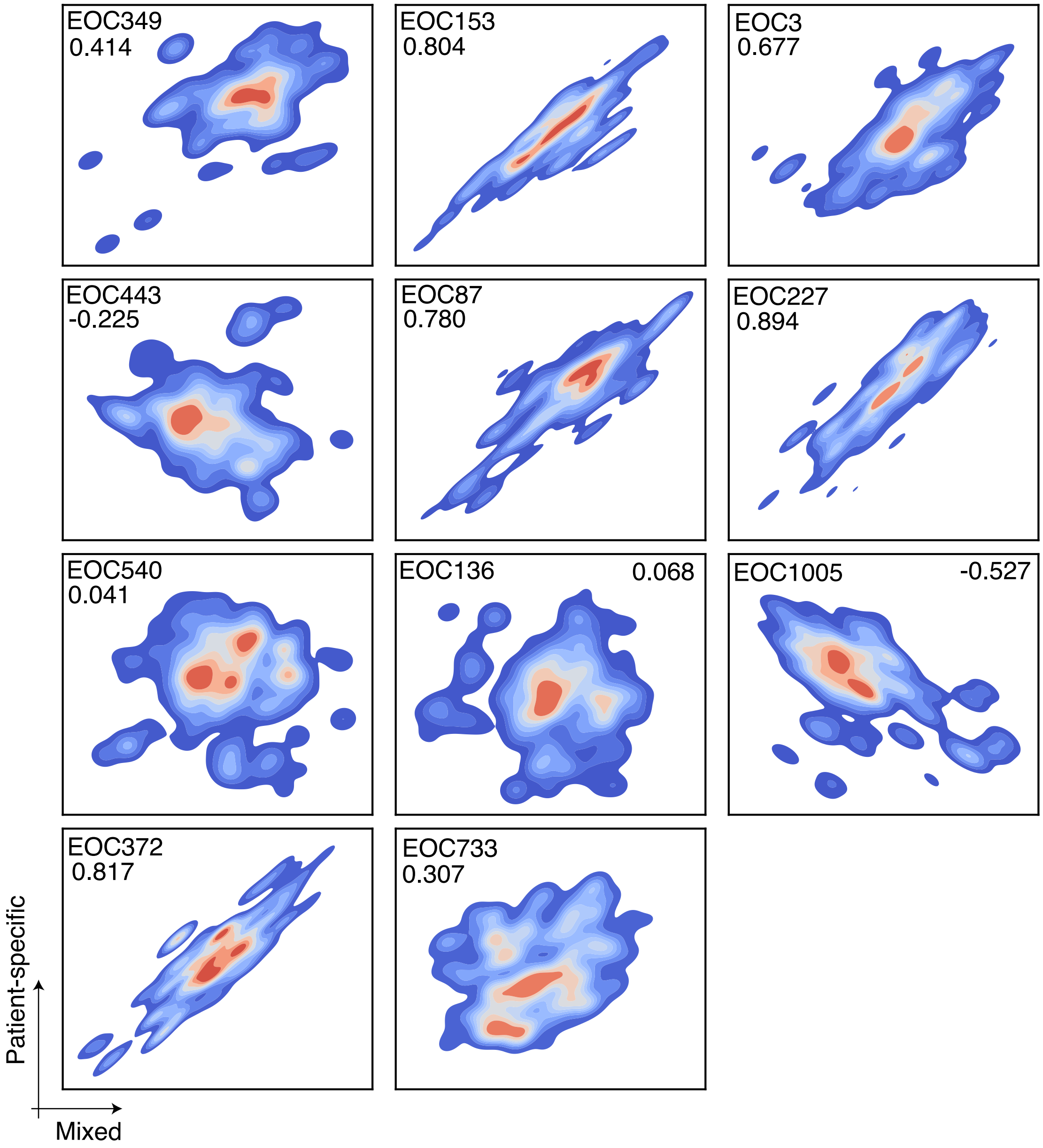

**Fig. S28**: Patient-wise contour plots of CNV profiles generated using `copyKAT` from patient-specific and mixed representation of ovarian cancer data (value is Spearman's correlation).

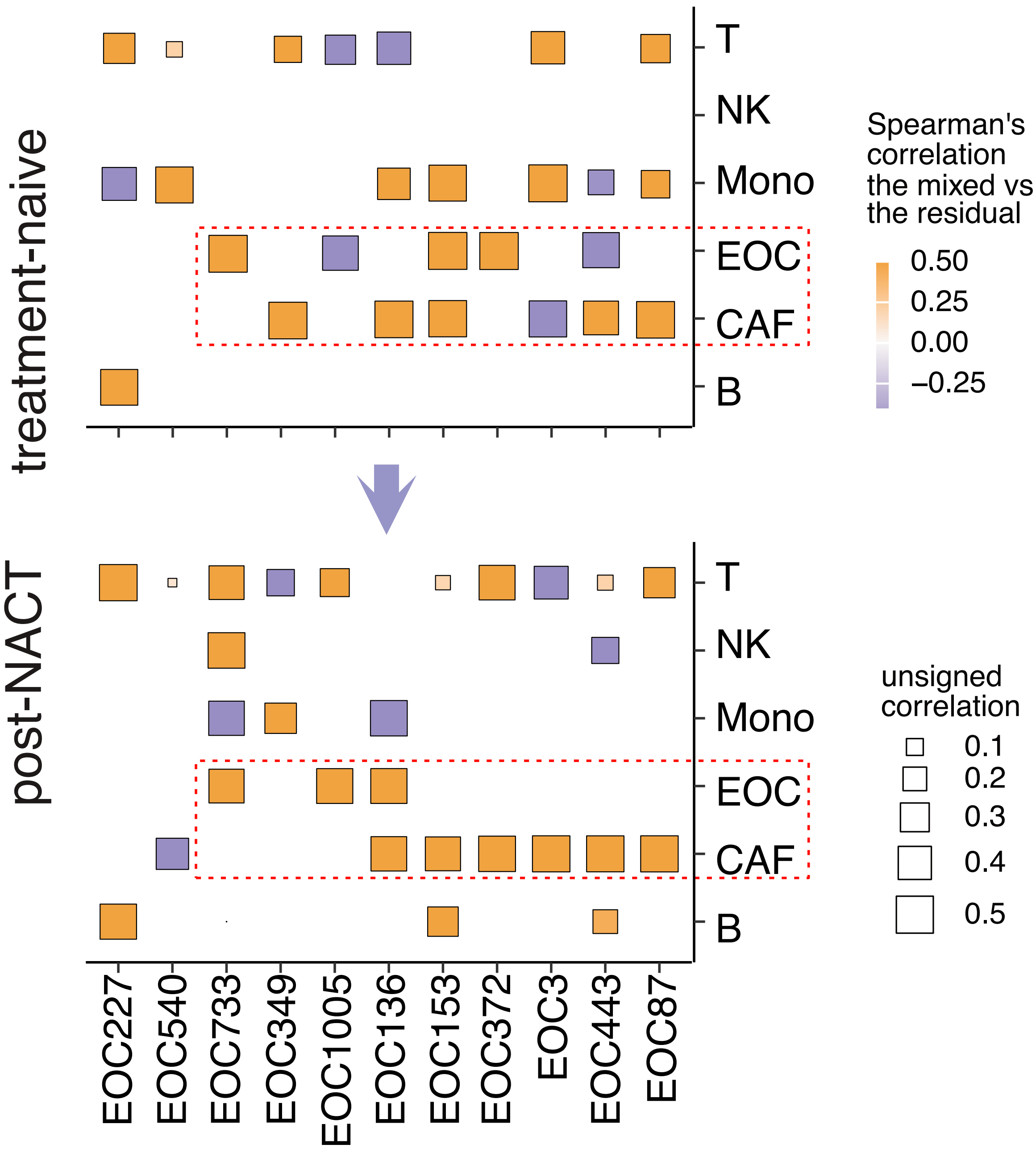

**Fig. S29**: Heatmap of correlation between CNV profiles from patient-specific and mixed representations for different cell types at treatment-naive and post-NACT stages.
